# Aberrant fast spiking interneuronal activity precedes seizure transitions in humans

**DOI:** 10.1101/2024.01.26.24301821

**Authors:** Edward M. Merricks, Sarita S. Deshpande, Alexander H. Agopyan-Miu, Elliot H. Smith, Emily D. Schlafly, Guy M. McKhann, Robert R. Goodman, Sameer A. Sheth, Bradley Greger, Paul A. House, Emad N. Eskandar, Joseph R. Madsen, Sydney S. Cash, Andrew J. Trevelyan, Wim van Drongelen, Catherine A. Schevon

## Abstract

There is active debate regarding how GABAergic function changes during seizure initiation and propagation, and whether interneuronal activity drives or impedes the pathophysiology. Here, we track cell-type specific firing during spontaneous human seizures to identify neocortical mechanisms of inhibitory failure. Fast-spiking interneuron activity was maximal over 1 second before equivalent excitatory increases, and showed transitions to out-of-phase firing prior to local tissue becoming incorporated into the seizure-driving territory. Using computational modeling, we linked this observation to transient saturation block as a precursor to seizure invasion, as supported by multiple lines of evidence in the patient data. We propose that *transient* blocking of inhibitory firing due to selective fast-spiking interneuron saturation—resulting from intense excitatory synaptic drive—is a novel mechanism that contributes to inhibitory failure, allowing seizure propagation.

## Introduction

Anti-seizure medication is unsuccessful in close to a third of epilepsy patients, resulting in uncontrolled seizures in nearly 20 million people worldwide^1^. Understanding the underlying mechanisms of seizure generation and subsequent spread through cortex is vitally important for efforts to improve management of epilepsy. At its most basic level, seizures are sometimes characterized in terms of an imbalance between excitatory and inhibitory activity, but recent research has highlighted a more nuanced view^2,3^, focusing instead on positive and negative feedbacks within the network^4,5^. Particular attention has focused on whether usually inhibitory interneurons may paradoxically excite their postsynaptic targets under certain conditions^6–11^ and even initiate the seizures themselves, possibly through pathologically re-wired networks^2,12,13^ or as a result of rebound excitation after synchronous inhibitory activity^14–16^.

A longstanding hallmark of epileptic physiology is that of surround inhibition, in which the intense synaptic drive arising from a seizure nearby is restrained by feedforward inhibitory activity in the surrounding “penumbral” territory^17–20^. Direct evidence for this mechanism has been well-established in animal models^21–25^, and neuronal firing patterns indicative of its presence have been shown in human recordings, albeit without direct assay of inhibition^26–28^.

As a result, both increases^14–16,29–31^ and decreases^32–34^ in interneuronal firing have been theorized to underlie seizure onset and spread. A dynamic interplay between the successful inhibitory restraint and its subsequent failure, allowing the seizure to propagate, may suggest that both mechanisms coexist to varying degrees in naturally occurring seizures^2,35^. However, the exact mechanism by which the inhibitory surround fails remains unresolved, impeding the identification of potential therapies. This collapse could be mediated either by the interneurons themselves becoming overwhelmed, or due to their own activity overwhelming the postsynaptic cells. For example, depolarization block could result in interneurons ceasing to fire^24,32,36,37^. Alternatively, increases in postsynaptic intracellular chloride ion concentration^2,7,9,38^ or extracellular potassium ion concentration^2,39,40^ could result in weakened interneuronal efficacy or even result in excitatory effects. Furthermore, recent animal work has posited that the failure may arise due to reorganization of activity in specific inhibitory cell-types rather than either of the aforementioned pre- and post-synaptic alterations^41^.

Experimental models have helped considerably in teasing apart these mechanisms, however there are likely many paths to ictogenesis, and being able to elicit seizures experimentally does not necessarily equate to uncovering specific mechanisms during unprovoked human focal seizures. In particular, the role of interneuronal activity during the pathological spread of naturally occurring epileptic activity remains a complex and open question. To this end, here we analyzed microelectrode recordings during spontaneous neocortical seizures in patients undergoing presurgical monitoring for focal epilepsy, enabling examination of the spatiotemporal activity patterns of populations of individual neurons, grouped by their putative cell types, and contextualized their firing patterns with a computational model.

## Results

### Neocortical seizure involvement is defined by transient increases in interneuronal firing rate

Spontaneous seizures captured on microelectrode array (MEA) recordings from thirteen patients were analyzed, up to a maximum of three per patient (Table S1). MEAs were arranged in a 10 x 10 grid over 4 x 4 mm with either 1- or 1.5-mm length electrodes, and implanted into the presumptive seizure onset zone in neocortical gyri. 4,222 single units were isolated across 33 seizures (mean ± SD per seizure: 127.9 ± 67.7), with 11.98% of units being probabilistically subclassified as inhibitory interneurons^42^ (see Methods). The numbers of cells belonging to each classification was robust, with the putative inhibitory population dropping only by 3.84% when restricting to a high confidence threshold of 95% (Fig. 1).

**Figure 1.**
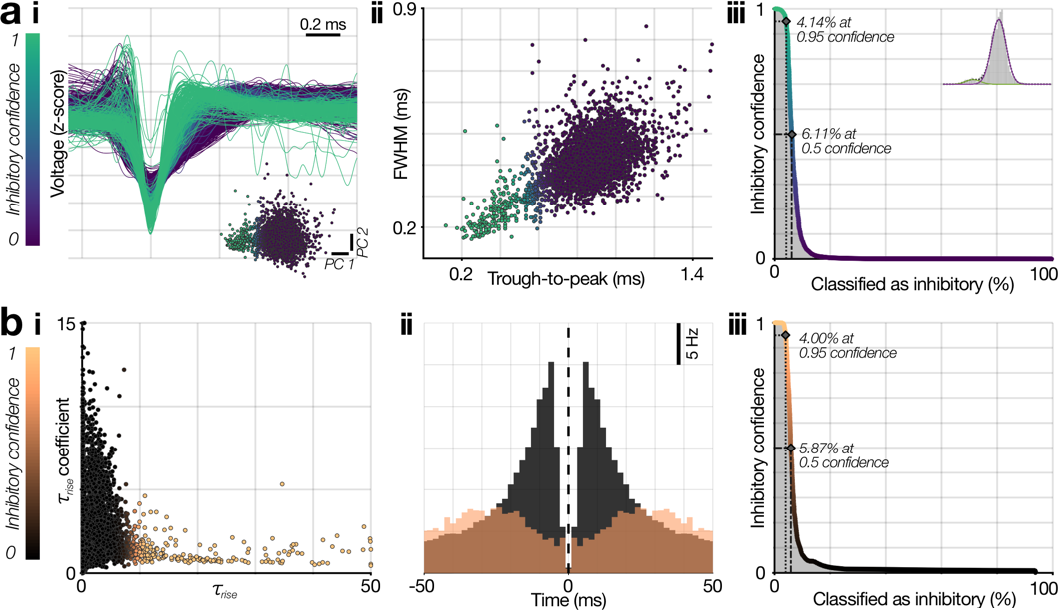
Cell-type subclassification of single-units. (**a**) Probabilistic subclassification of fast-spiking (FS) interneurons. (***i***) Mean wideband waveforms from each single-unit, color-coded by its confidence of being classified as inhibitory based on a 2-component Gaussian mixture model fitted to these waveforms’ scores in PC space (inset; color-scale maintained throughout A). (***ii***) Confidences from this model showed the expected differences in spike duration in the putative FS interneuron population, and classification proved stable, with similar percentages of units being classified as FS interneurons across a wide range of confidence cutoffs (**iii**; inset: Fisher-linear discriminant projection of the 2-component Gaussian-mixture model). (**b**) Probabilistic subclassification of excitatory cells. (***i***) The autocorrelograms for each unit that was not classified as a putative FS cell were fitted with a set of exponentials to calculate τ_rise_ (Petersen et al., 2020;^81^ see Methods), which were then fitted with a 2-component Gaussian mixture model to derive confidences of each unit being excitatory or inhibitory (black to copper color scale; maintained throughout **b**). (***ii***) Mean autocorrelograms for wide single-units with > 50% and ≤ 50% confidence of being inhibitory based on firing patterns (copper and black respectively). (***iii***) As for putative FS interneuron classification, the probabilistic classification of the remaining excitatory versus inhibitory population proved stable, with similar percentages of units being classified across a wide range of confidence cutoffs.

Clinical macro-electrodes are dominated by synaptic activity as opposed to local neuronal firing^43^, and so primarily show the *input* to the local tissue. As a result, ictal patterns on the EEG are not sufficient to determine if a region has been recruited into seizure-driving territory (the spatiotemporally dynamic ictal “core”) or if intact inhibition is successfully restraining the intense excitatory drive at that moment (“penumbra”)^22,26,44^. We have previously shown that a wavefront of continuous (“tonic”) firing occurs at the boundary when penumbral tissue becomes recruited^26,28,45–47^ and that individual neurons undergo action potential waveform alterations upon recruitment^28,42,44^. This activity is spatially restricted, with these firing patterns coexisting within seizures, marking the propagation of ictal activity (Fig. 2). A sustained, significant increase in firing rate therefore defined the moment of local recruitment at individual electrodes^45^ (see Methods for description of quantitative criteria), and ictal recordings without this signature activity were classified as unrecruited.

**Figure 2.**
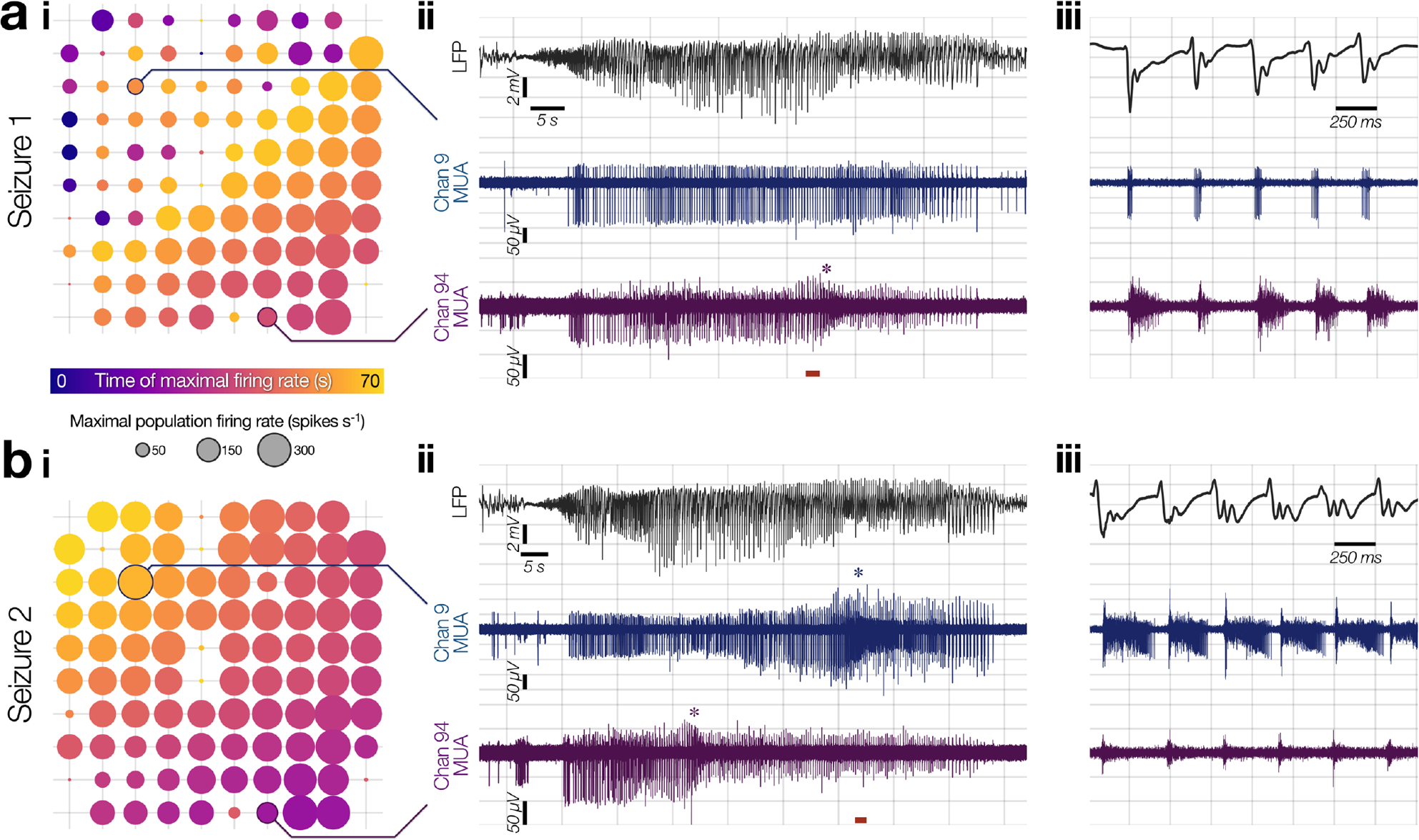
Spatiotemporal dynamics of ictal recruitment across the MEA. (**a, b**) Consecutive seizures captured 5.5 hours apart (Patient 7, seizures 1 & 2). (***i***) Firing rate spatiotemporal patterns during the seizure: each circle corresponds to a single MEA electrode, where its size denotes the maximal population firing rate that occurred at that electrode during the seizure, and its color shows when that maximal firing occurred. Note the maintained propagation direction (color-scale, running from bottom right to top left) across both seizures, but the sharp diagonal in the maximal firing rate during the first seizure, showing the outer boundary of ictal recruitment before the seizure terminated in seizure 1 while it successfully propagated across the whole array in seizure 2. (***ii***) Mean LFP (2–50 Hz bandpass) from the whole MEA (top) with paired MUA (300 Hz–5 kHz bandpass) from example channels highlighted in ***i*** (below). Asterisks show the timing of local recruitment at that electrode (i.e., when local neurons become involved in *driving* the seizure rather than simply being impacted by the excitatory barrages from elsewhere, see Methods): note the absence of recruitment in channel 9 in seizure 1, and the temporal relationship between recruitment in channels 9 and 94 in seizure 2, in alignment with the extent of ictal propagation seen in ***i***. The epoch between “global” seizure onset (earliest evidence of ictal activity in the LFP) and local recruitment is denoted “pre-recruitment” for that channel. (***iii***) Enlargement of the short epochs marked with red bars in ***ii***, highlighting, in **a**, a moment when channel 94 is becoming recruited to the seizure while channel 9 (∼3.2 mm away) remains unrecruited in seizure 1, and in **b**, when channel 94 has been recruited and the seizure core has now reached channel 9, in seizure 2.

Note “local” recruitment timing differs from the first EEG changes (“global” seizure onset) since it takes into account how slowly seizures propagate, largely due to delays that can be attributed to a powerful inhibitory restraint. For each seizure, therefore, the timing of *global* onset is a single time point—when the seizure has started but most tissue remains penumbral—while the timing of *local* recruitment varies across sites (Fig. 3). The transition to pathological activity must occur between these points, with the timing of local recruitment being critical. However, human studies to date have focused on inhibitory activity aligned to global onset, with local recruitment remaining uncertain. We therefore sought to characterize firing patterns with respect to the moment of local recruitment.

**Figure 3.**
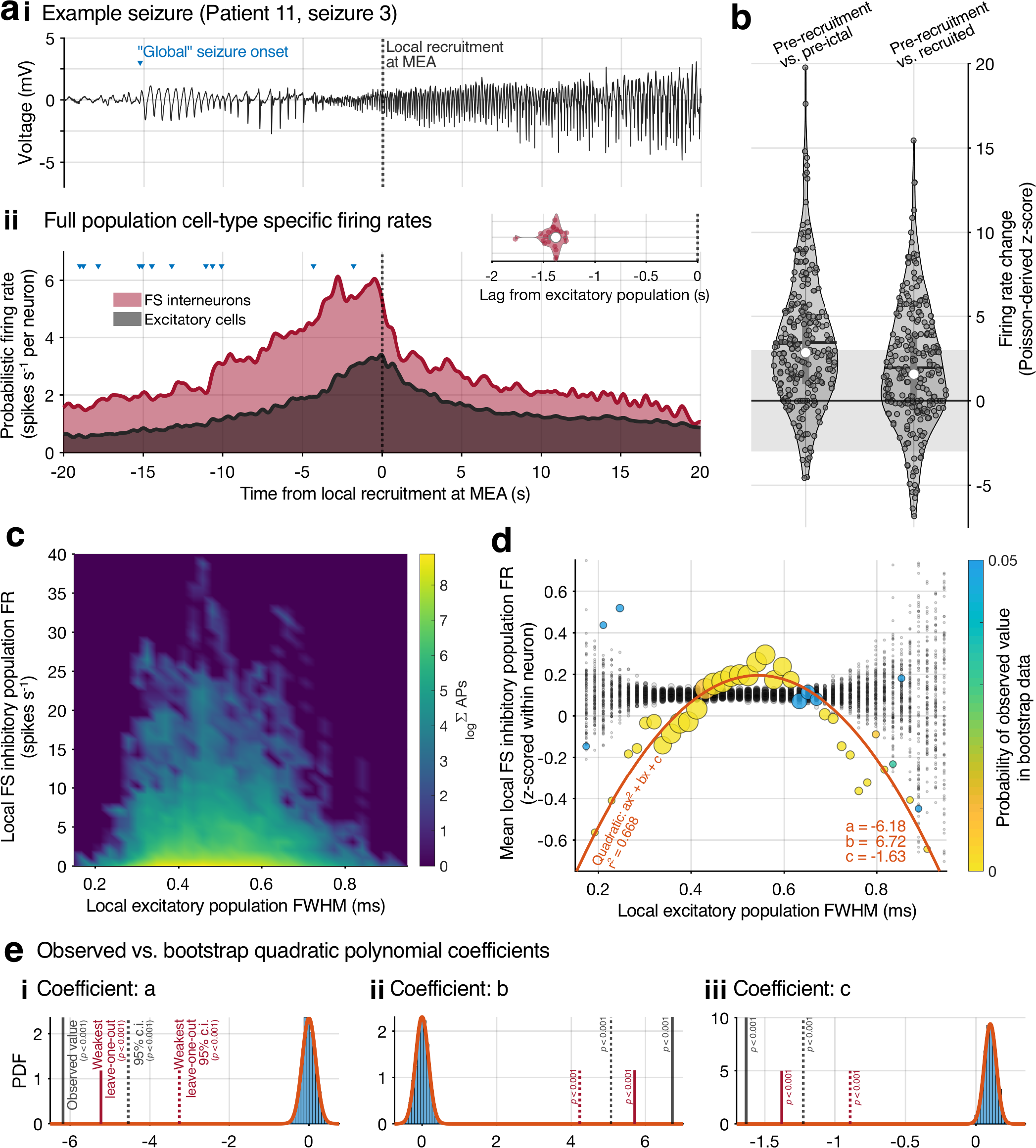
Firing rate patterns with respect to local ictal recruitment. (**a**) LFP (2–50 Hz bandpass) from an example seizure, showing global onset (blue triangle) and the average moment of local recruitment at the MEA (dotted line, ***i***), with paired population firing rate data from all seizures aligned to the average moment of local recruitment at the MEA (***ii***), for fast-spiking (FS) interneurons and excitatory cells (red and black respectively). Blue triangles show global onset for individual seizures. Note the increase in FS interneuronal preceding local recruitment, when excitatory firing peaks. This > 1 second precession is stable across a leave-one-out analysis (lag at maximal cross correlation between populations, inset). (**b**) Poisson-derived z-scored firing rate changes for FS interneurons between the pre-recruitment epoch and the pre-ictal 30 seconds (left) and the post-recruitment epoch (right; in each, mean: black line; median: white dots; individual FS interneurons: gray dots). (**c**) Heatmap of FS interneuron firing rates across all seizures as a function of the excitatory population’s spike full-width at half maxima (FWHM) at the same electrode—a metric of local ictal recruitment—showing an increase followed by decrease in population firing rates. (**d**) Equivalent to **c**, but with normalized FS interneuron firing rates (colored dots; z-scored within each neuron) and bootstrap results from repeat runs while shuffling the order of the data within each neuron to create null distributions (black dots). Diameter of each dot corresponds to the total number of observations in the original data, which was maintained during the bootstrap analysis. Color scale shows probability of the observed data in the null distributions. Orange line: polynomial fit to the observed data. (**e**) Coefficients from the polynomial fit to: the observed data (solid black lines; 95% confidence in dotted black lines); the weakest results from the observed data during a leave-one-out analysis (red lines); and the distribution of values and a gaussian fit for the bootstrap data (blue histogram and orange line respectively). *P* < 0.001 in all observed scenarios.

Seizures that included evidence of recruitment at the recorded location (27 seizures from 11 patients; Table S1) showed a stereotyped, transient increase in inhibitory firing after global seizure onset and prior to the excitatory increase at local recruitment (the “*pre-recruitment*” epoch; Fig. 3a). Across the full population, this inhibitory activity occurred 1.40 seconds before excitatory increases (cross-correlation analysis, *r*-squared = 0.984; Fig. 3a ii *inset*) and was similar across all seizures (leave-one-out analysis, mean ± SD: lag = 1.41 ± 0.098 s; *r*-squared = 0.983 ± 0.0014). For individual FS interneurons, the transient increase in firing rate during pre-recruitment was larger than pre-ictal levels in the majority of cells (Fig. 3b; pre-ictal mean ± SD: 0.86 ± 1.25 spikes s^−1^; pre-recruitment: 3.39 ± 4.19 spikes s^−1^; *P* = 9.09 x 10^−22^, Mann-Whitney U test; increases in 82.8% of FS interneurons) and similarly larger than firing post-recruitment until seizure termination (Fig. 3b; mean ± SD firing rate: 1.80 ± 3.11 spikes s^−1^; *P* = 1.09 x 10^−6^, Mann-Whitney U test; increases in 67.2% of FS interneurons). While increases were largest after global onset and prior to local recruitment, recruitment was not associated with reductions in inhibitory firing, with increases over baseline in 193 of 248 of FS interneurons (77.8%) and only 8 (3.2%) showing complete cessation of firing (6 in patient 8, seizure 1, 1 in patient 5, seizure 2 and 1 in patient 9, seizure 1).

Due to this firing rate increase after global onset but *before* local recruitment, we hypothesized that it was primarily caused by feedforward inhibition from the approaching ictal wavefront^22,23,26^, which *in vitro* animal models have suggested is mediated first by parvalbumin-positive (PV^+^) interneurons^25,41^, which have been shown to primarily consist of fast-spiking (FS; Fig. 1a) cells^48,49^. We therefore analyzed the firing response of this FS population as a function of spike full-width at half maximum (FWHM) of all other cells recorded at the same electrode (Fig. 3c, d). We previously showed how spike FWHM is a suitable indicator for ictal recruitment at the level of single neurons^28^; it therefore provides the opportunity to quantify the degree to which a region has been impacted, continuously.

In keeping with the model of feedforward inhibition ahead of the ictal wavefront that subsequently collapses, FS interneuronal firing rates showed a clear “inverted U” response as the excitatory cells surrounding them displayed increasing FWHM values (Fig. 3c). This response of increasing inhibitory firing rates followed by a collapse as the surrounding tissue underwent larger waveform alterations, was robustly modeled with a simple quadratic fit (*r*-square = 0.668; Fig. 3d). Comparison to a null dataset— created by shuffling the firing rate values at random 1,000 times and re-fitting the data—confirmed the negative quadratic was unlikely to arise by chance (*P* < 0.001 in each coefficient, Holm-Bonferroni corrected; Fig. 3e). Similarly, a leave-one-out analysis confirmed the response was stable across seizures, with the most conservative estimates still significantly removed from the null dataset (*P* < 0.001 in each; Fig. 3e).

This firing pattern can be seen clearly in patient 7’s two consecutive seizures, where the first propagated across only half the MEA, before terminating, and the second successfully propagated across the entire MEA (Fig. 2). These two contrasting seizures thus provided an opportunity to compare neuronal behavior first, between recruited versus unrecruited tissue simultaneously recorded on the same array in the first seizure, and second, of the same cortical territory being unrecruited and then recruited across subsequent seizures, thereby demonstrating the variable extent of ictal recruitment from one seizure to the next. These recordings emphasize the spatial relationship of feedforward inhibition ahead of the propagating ictal wavefront with ramping up of each FS unit’s firing starting at global onset, and increasing as the seizure propagates closer, before subsiding as the local tissue becomes recruited (Supplementary Movies 1 & 2). This in itself, of course, is not indicative of whether FS interneuron firing rate reduction is causative of local recruitment, or a result of the pathological activity. To further explore this relationship, we turned to analysis of rhythmic onset seizures specifically to be able to relate cell-type specific neuronal firing to repetitive ictal discharges^50^.

### Out-of-phase FS interneuron activity in rhythmic onset seizures

A subset of 18 seizures from eight patients showed rhythmic onset patterns (> 2.5 SD increase of power in either delta, theta or alpha bands before ictal recruitment; Table S1). These seizures allowed the opportunity to analyze inhibitory firing patterns from ictal onset through local recruitment with respect to the dominant ictal rhythm, while the higher frequencies in low-voltage fast onsets preclude correlating neuronal firing to ictal discharges. Prior to recruitment, ictal discharges were accompanied with entrained neuronal firing across both excitatory and inhibitory cells (*P* < 0.001 in each, across all seizures, Rayleigh *z*-test; Fig. 4a).

**Figure 4.**
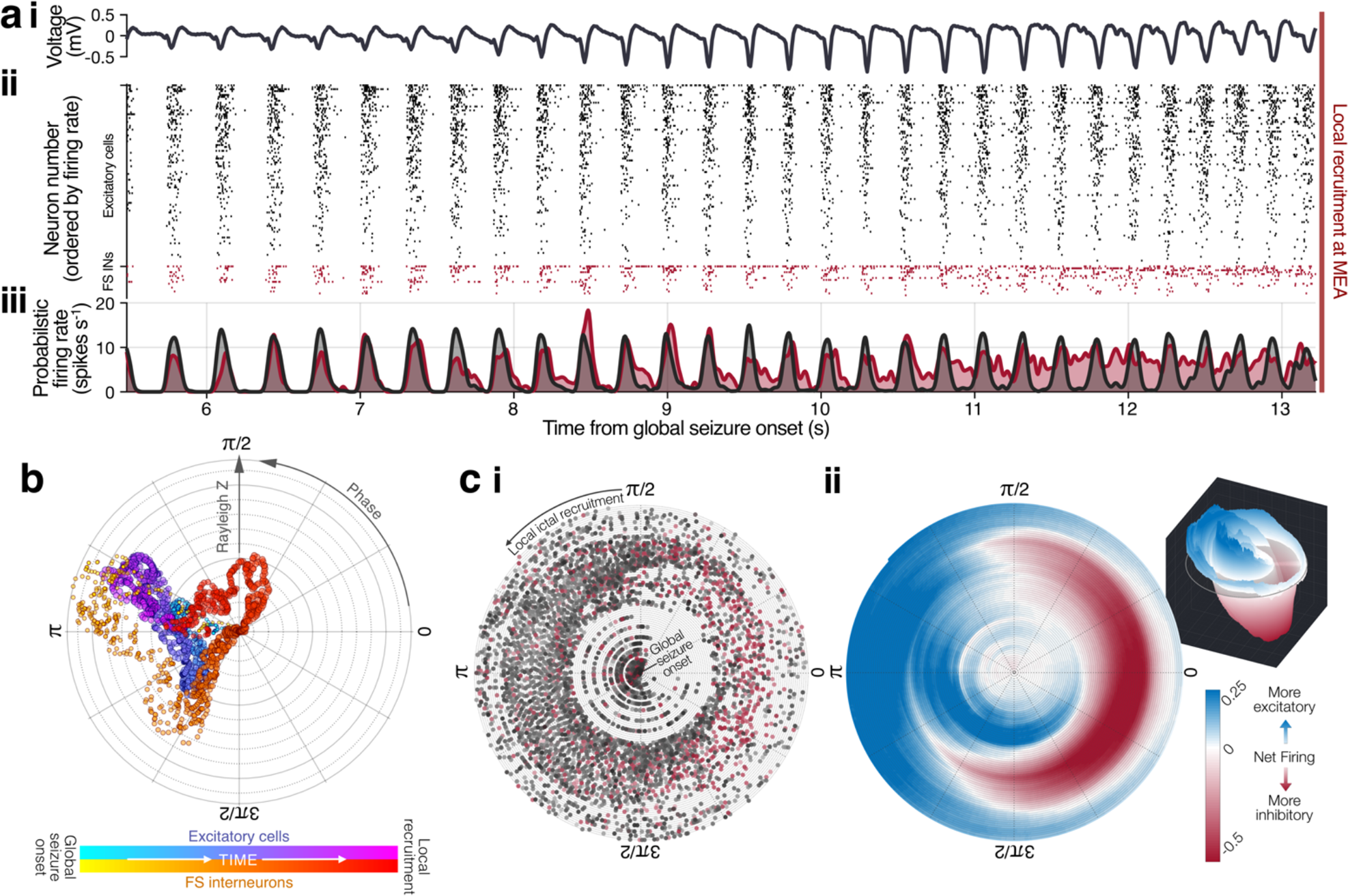
Cell-type specific differences in the lead-up to local ictal recruitment. (**a**) Activity in the lead-up to local recruitment for an example rhythmic onset seizure (Patient 3, seizure 1). (***i***–***iii***) Mean LFP across the array (2–50 Hz bandpass), raster plot for all isolated neurons (red: FS interneurons; black: all other cells), and gaussian-convolved probabilistic firing rate for the FS interneuron and excitatory cell populations (red and black respectively). (**b**) Mean phase entrainment through time (2 second bins, advancing in 10 millisecond increments) for FS interneuron and excitatory cell populations in the pre-recruitment epoch for the same seizure shown in **a**. Angles show mean phase direction, radius shows level of instantaneous entrainment to the dominant ictal rhythm (Rayleigh z value, normalized within cell-type), dot diameters are scaled by the total number of action potentials contributing to that bin. Note the transition of the FS interneuron population to anti-phase firing before recruitment. (**c**) Same epoch as **b**, showing all action potentials across the population through time on a spiral, revealing the temporal evolution of the phase angles (***i***). Each dot is one action potential (red: FS interneuron; black: excitatory cell; saturation is the confidence that action potential arose from its assigned neuron). The location of each dot represents both the progression of time as the radius of the spiral expands, and the instantaneous phase of the dominant ictal rhythm at that time as the angle. (***ii***) Calculating the gaussian-convolved firing rates for the two populations across the spiral (see Methods) and subtracting the inhibitory from the excitatory gives an instantaneous estimate of inhibitory versus excitatory firing (color-scale; three-dimensional view inset), thereby characterizing the spatiotemporal dynamics of the neuronal firing with respect to the dominant ictal rhythm.

Ahead of the invasion of the ictal wavefront, however, FS interneuronal bursts were seen to slip out-of-phase both from the ECoG signal and from the remainder of the neuronal population (Fig. 4b), suggesting a pathological mechanism altering firing burst patterns as opposed to a simple disruption to interneuronal firing. In the 5 seconds before recruitment there was an increase in the circular distance between the FS interneuron and pyramidal cell population phases (*P* = 2.63 x 10^−83^, Mann-Whitney U test). This shift in entrainment was echoed at the individual seizure level, with significant differences in the cell-type specific phase distributions in each seizure (*P* < 0.05 in each, range: *P* = 0.002–0.018, Holm-Bonferroni corrected Kuiper test).

To quantify and to visualize the spatiotemporal dynamics of this cell-type specific transformation, we analyzed firing patterns with respect both to instantaneous phase and ictal time simultaneously (Fig. 4c). Each rhythmic onset seizure showed periods of anti-phase FS interneuron-dominant firing prior to local ictal recruitment (Fig. 5). At the population level, inhibitory firing lagged the peak of the excitatory firing (Fig. 5a; *P* < 0.001, Kuiper-test). While the exact phase of this inhibitory firing with respect to the dominant ictal rhythm differed across patients (circular mean ± SD: 239.0° ± 74.44°), it was similar between seizures within each patient, with no measurable deviation from a von Mises distribution of equal concentration centered on 0° (Fig. 5b; von Mises fit to observed data: µ = −9.77°, κ = 0.65; *P* = 0.987. Similarly, there was no difference for a wide range of von Mises concentration parameters: *P* > 0.05 for 0 ≤ κ < 1.5). Each seizure was characterized by a relationship between the inhibitory firing and both the phase of the dominant ictal rhythm (angle in Fig. 5c) and the progression of time (radius in Fig. 5c; *P* < 1 x 10^−8^ in each, Holm-Bonferroni corrected multi-linear regression F-test). Thus, altered timing of interneuron burst firing prior to local recruitment was ubiquitous among the 8 patients and 18 seizures analyzed, and the firing pattern alteration was stereotypical for each patient.

**Figure 5.**
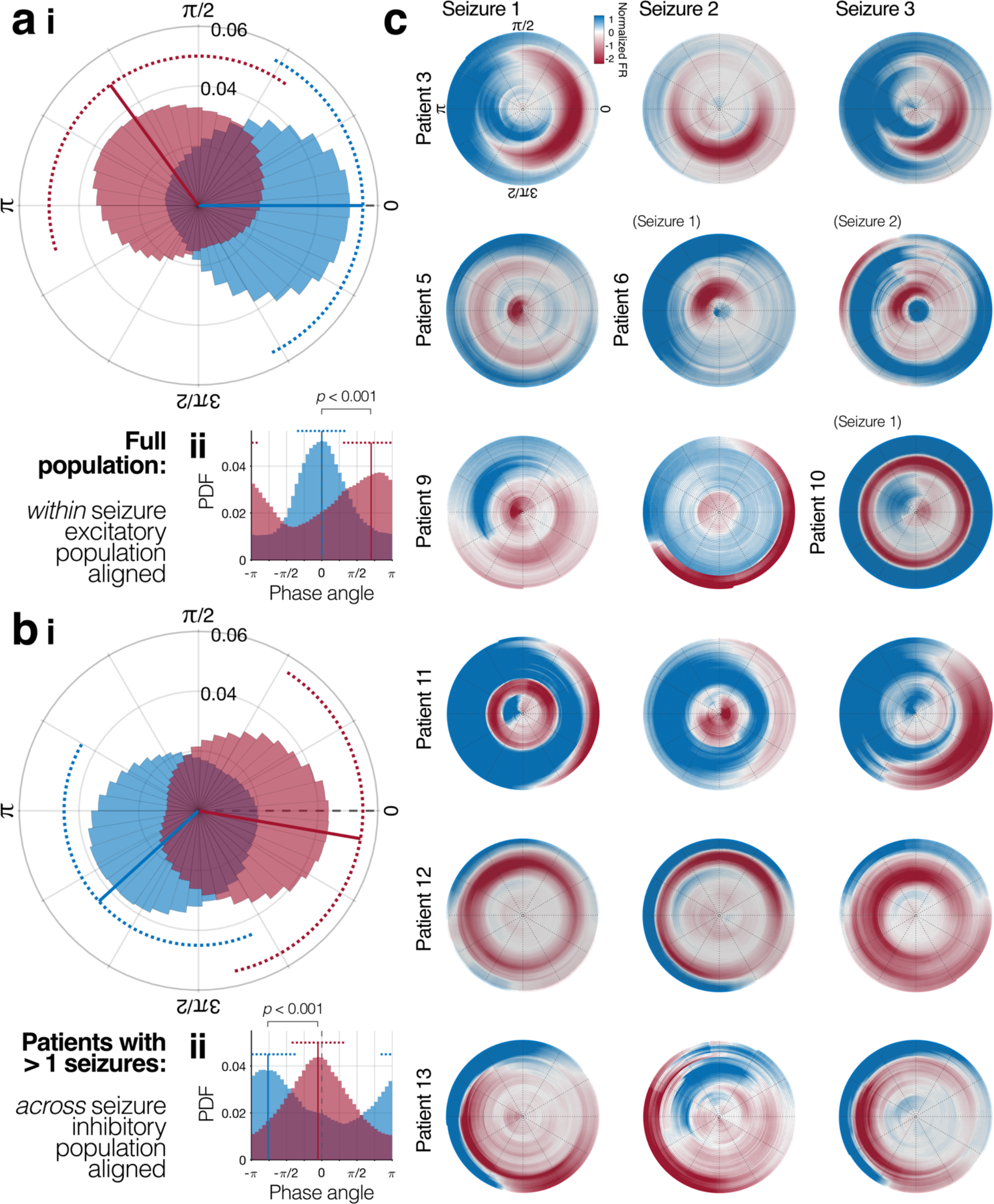
Out-of-phase inhibitory firing is a defining feature of rhythmic onset seizures. (**a**)(***i***) Phase angles of the dominant ictal rhythm for all action potentials (red: FS interneurons; blue: excitatory cells) between global seizure onset and local recruitment at the MEA, across all rhythmic onset seizures, normalized to the circular mean of that seizure’s excitatory population. Solid and dotted lines show circular mean and standard-deviation of each population respectively. (***ii***) Same data plotted in cartesian space. (**b**) Same as for **a**, but restricted to patients with multiple seizures (*n* = 16 seizures, 6 patients), and aligned instead to the circular mean angle of FS inhibitory firing across all seizures within that patient, revealing stable phase preferences across seizures within patient (pairwise distance of inhibitory phase angles within patients: 40.15° ± 29.64°; pairwise distance of inhibitory vs. excitatory phase angles within patients: 98.12° ± 52.07°; *P* = 9.39 x 10-5, unpaired t-test). (**c**) Cell-type specific firing phase/time plots as per Fig. 4c for each rhythmic onset seizure.

### Transient depolarization block in FS interneurons gives way to ictal recruitment

The change in phase angle in the 5 seconds leading into recruitment was accompanied by a shift to tonic firing exclusive to the FS interneuron population: while entrainment increased in pyramidal cells (*P* < 0.001, Rayleigh *z*-test), it decreased in FS interneurons (*P* = 0.031, Rayleigh *z*-test). We hypothesized that this anti-phase FS interneuronal activity preceding local ictal recruitment is driven by the approaching seizure’s ictal wavefront, which generates powerful excitatory synaptic currents that spread orders of magnitude faster than the speed of the ictal wavefront itself^26,46,51,52^. Under this hypothesis, PV^+^ interneurons would be impacted prior to other cell types due to the combination of feedforward synaptic wiring^53^ and their relatively small somata^37^. A defining feature of neurons that have been recruited to seizures is the paroxysmal depolarizing shift (PDS), which results in a loss of action potential amplitude^28,54–57^.

Analysis of the action potentials for individual putative FS interneurons revealed reduction in amplitude within the out-of-phase burst between ictal discharges, with a progressive loss of amplitude prior to local ictal recruitment (Fig. 6a). Averaging across all FS interneurons within a seizure, separated by consecutive discharges, revealed that not only did the mean amplitude decrease prior to recruitment, but decreased within each burst period, followed by a progressively smaller recovery in amplitude between discharges each time (Fig. 6b). Expanding this analysis across all rhythmic onset seizures revealed the same observations held true at the population level (Fig. 6c; z-scored amplitude change: −0.52 s^−1^ and −8.5 x 10^−3^ s^−1^ for intra- and inter-burst respectively; *P* < 0.001 in each, linear regression F-test).

**Figure 6.**
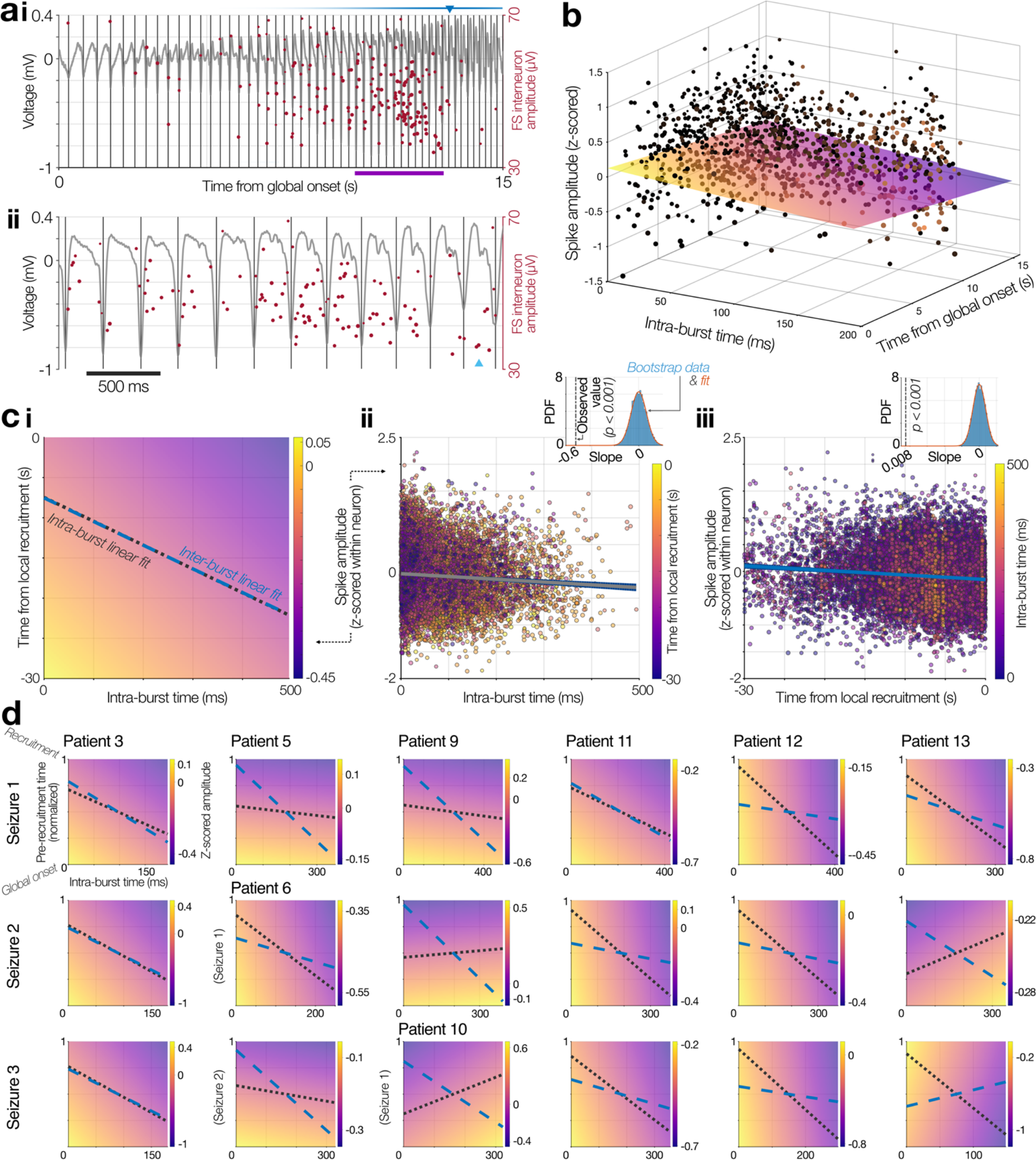
Fast-spiking interneurons undergo repetitive, transient loss of amplitude across discharges. (**a**)(***i***) Same seizure as Fig. 4a, showing an example FS interneuron’s action potential amplitudes through time (red dots, each scaled by confidence of match to that neuron). Black lines show the trough of each ictal discharge in the mean LFP across the MEA (gray trace). Blue triangle: mean local recruitment at the MEA; blue bar: probability density function for the distribution of local recruitment times across the MEA. (***ii***) Enlargement of the epoch marked in purple in ***i***. Cyan triangle: local recruitment at the shown FS interneuron’s channel. (**b**) Action potential amplitudes from all FS interneurons from the same seizure in **a**, with action potentials from each discharge plotted on the x-axis (intra-burst time), aligned to when that discharge started with respect to global seizure onset (y-axis). Dot color represents the ordinal number for the action potential within its neuron during that discharge, and diameter shows confidence it arose from its assigned neuron. Colored plane shows a 2D linear regression fit to these spike amplitudes. (**c**) Equivalent linear regression fits for the full population of FS interneurons across all rhythmic onset seizures, showing the 2D plane, the intra-burst data, and the continuous data (***i***–***iii*** respectively). Black dotted line and blue dashed line in ***i*** show the linear fits for intra- and inter-burst data respectively, on the same scale as the color bar. Insets in ***ii*** & ***iii*** show the observed coefficient versus bootstrap data from repeat runs while shuffling the temporal order of action potentials, revealing a significant reduction in amplitude both within discharge and through time. (**d**) 2D linear regressions and accompanying slope fits as per **c *i***, for each rhythmic onset seizure.

This pattern of loss of amplitude followed by decreasing levels of recovery between each burst was remarkably stable across all rhythmic onset seizures (Fig. 6d). On average, intra-burst loss of amplitude was −1.26 ± 1.74 SD s^−1^, with 15 of 18 seizures (83.3%) showing a downward average trajectory across all FS interneurons through time. For individual FS interneurons, intra-burst loss of amplitude was −21.24 ± 35.11 µV ms^−1^. These trajectories were a larger loss of amplitude than expected by chance, as compared to a bootstrapped null distribution derived from shuffling the temporal order of spikes 10,000 times (*P* = 1.48 x 10^−16^).

Inter-burst amplitudes also decreased prior to local ictal recruitment (−1.49 x 10^−2^ ± 1.79 x 10^−2^ SD s^−1^), with 17 of 18 seizures (94.4%) showing progressive pre-recruitment reductions in average FS interneuron amplitudes (Fig. 6d). Across the whole population, these inter-burst amplitude trajectories were a larger loss of amplitude than expected by chance, again comparing to a bootstrapped null distribution from shuffled spike orders, repeated 10,000 times (*P* = 4.44 x 10^−31^). We therefore hypothesized that these repetitive amplitude alterations are indicative of repeating, short-duration depolarization-inactivated action potentials^55,58,59^ due to the large excitatory barrage onto the PV^+^ interneurons.

### Out-of-phase firing and transient depolarization block of fast-spiking interneurons is inherent to a Hodgkin-Huxley model

The observed intra- and inter-burst amplitude trajectories in the FS population are in keeping with the hypothesis of repeated, brief depolarization blocks that eventually overwhelm their inhibitory restraint allowing the seizure to propagate into the local tissue. Given that patient recordings do not allow for experimental probing of the cause of these observed neuronal behaviors, we instead developed a computational model reproducing the effect, to test the hypothesis that the observed out-of-phase firing is an inherent result of the cell-type specific size and dynamics when excitatory input increases across the population.

Using a 10-neuron cortical model characterized by Hodgkin-Huxley type dynamics^58^, comprised of 20% inhibitory and 80% excitatory cells with cell-type specific activity profiles^60^ (Fig. 7a, b; see Methods), we were able to reproduce markedly similar firing patterns to the observed human data when injected with an ictal rhythmic input (Fig. 7c–f; c.f. Fig. 4). At seizure onset, the inhibitory and excitatory cells burst synchronously in-phase (Fig. 7e), followed by a phase delay in inhibitory bursting activity, ultimately leading to out-of-phase inhibitory bursting (Fig. 7f). Afterwards, the excitatory population begins to enter tonic firing, echoing observations in human recordings (Supplementary Fig. 1). Intracellularly, we see that the inhibitory population is characterized by decreasing spike amplitudes within bursts (Fig. 7e), followed by decreasing spike amplitudes overall (Fig. 7f). This phenomenon is associated with reduced synaptic transmission^37^, and is indicative of the inhibitory population transiently entering and exiting neuronal saturation, leading to the phase shift and eventual out-of-phase bursting activity captured extracellularly and in the net firing rate through time (Fig. 7d).

**Figure 7.**
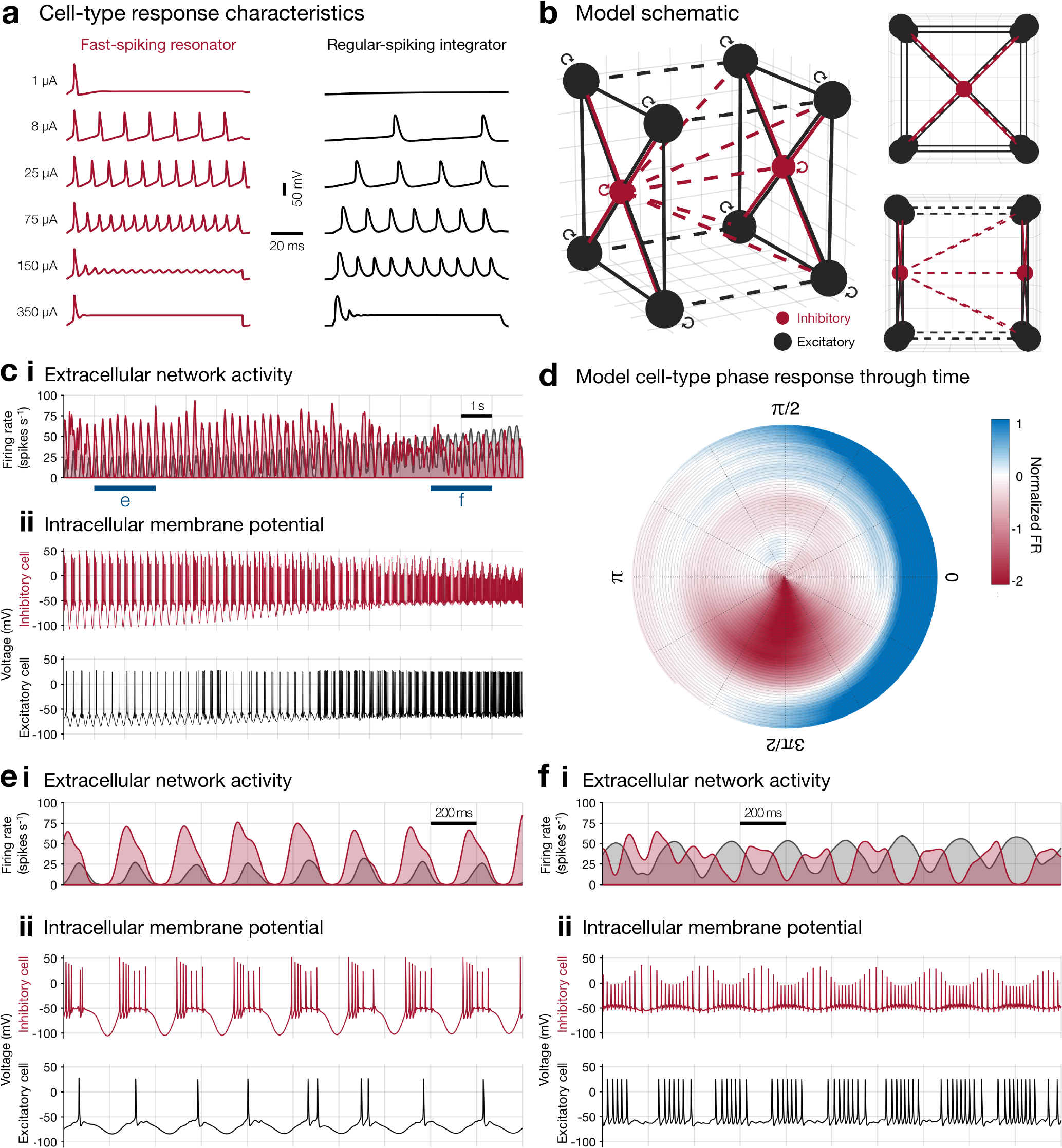
A Hodgkin-Huxley model of the ictal pre-recruitment period corroborates clinical patterns. (**a**) Cell type response characteristics for fast-spiking resonator type neurons (inhibitory; *red*) and regular-spiking integrator type neurons (excitatory; *black*). Fast-spiking resonator type neurons will saturate prior to regular-spiking integrator type neurons at lower input currents. (**b**) Model schematic of the 10-neuron model, with excitatory and inhibitory cells in a 4:1 ratio, including front and side view. (**c**) The extracellular network activity of the model corroborates the extracellular network activity of the patient data. At seizure onset, the inhibitory and excitatory populations burst in-phase. As the seizure progresses towards ictal recruitment, the inhibitory population begins to burst out-of-phase. This can be attributed to neuronal saturation of the inhibitory population, which can be observed by the decreasing spike amplitudes within bursts and overall in the intracellular membrane potential. (**d**) Cell-type specific firing phase/time plots as per Fig. 4c for the extracellularly detected spikes in the model. The instantaneous angle of the sine wave in the injected current provided the phase values since the local field potential is not calculated in the Hodgkin-Huxley model. (**e** & **f**) Magnifications for the marked epochs in (d), revealing the early in-phase activity of the inhibitory cells and later apparent out-of-phase activity respectively. Note the temporary intracellular loss of amplitude within discharges in the inhibitory cell in (e), and the further loss of amplitude resulting in loss of tracking in the extracellular data, including a lack of complete recovery between bursts as the modeled ictal wavefront approaches in (f).

These results offer a mechanistic basis to explain qualitatively the transition of inhibitory firing from in- to out-of-phase. In order, therefore, to make a quantitative comparison between the model and patient data, triple correlation—a method that fully characterizes spatiotemporal network activity^61^—was computed on the population spike data from the model and from a representative seizure (Patient 3, seizure 1; see Methods). Relative contributions of motif-classes quantitatively describe network activity at any given moment, and the prevalence of each motif-class through time was similar to the observed patient data (Fig. 8a; 0.0028 ≤ *p* ≤ 0.03, Benjamini-Hochberg-corrected).

**Figure 8.**
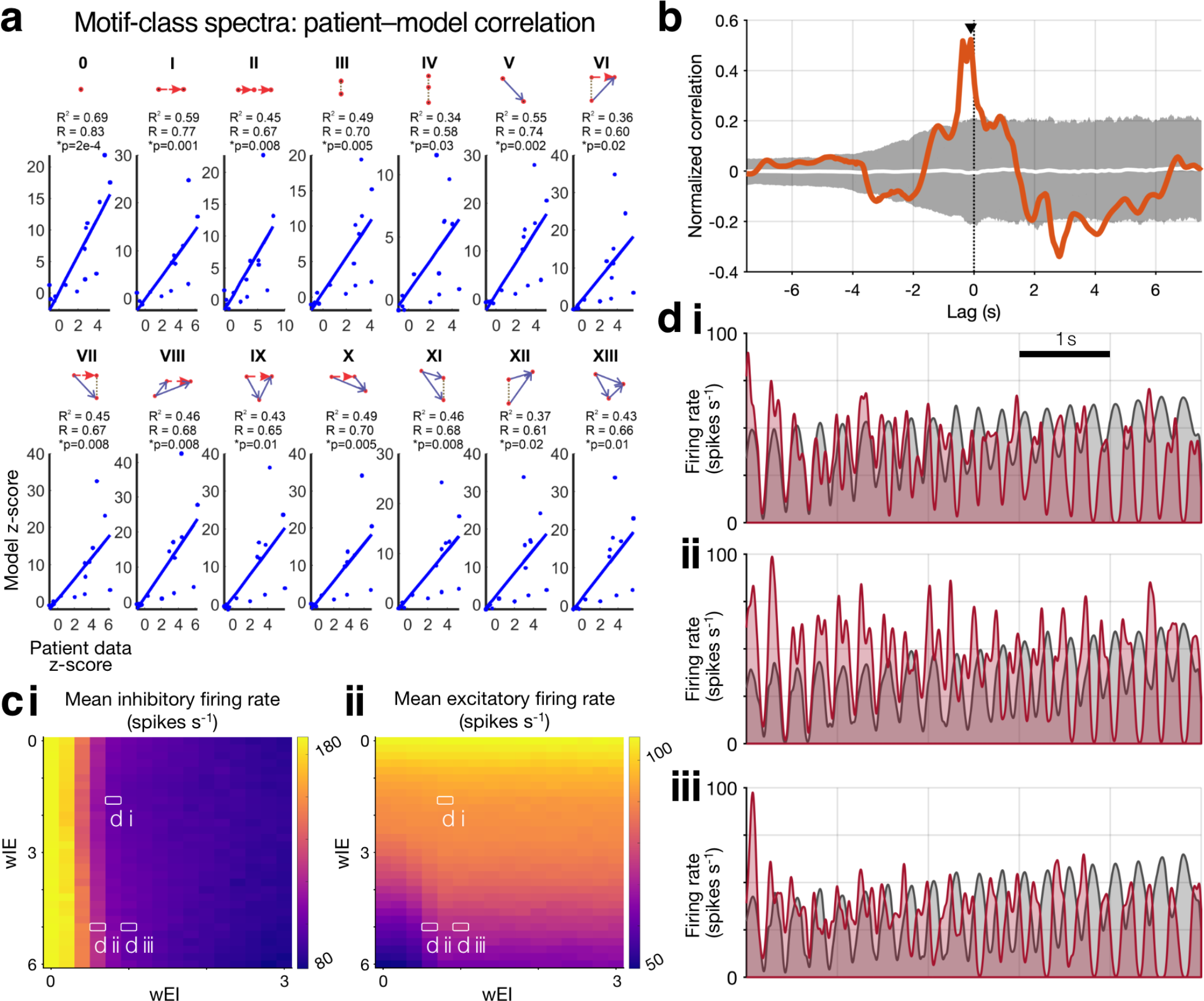
Quantitative network comparison and model parameter exploration. (**a**) The model output and observed patient data were significantly similar across all 14 motif-classes. One-second epochs were normalized to the values in each motif-class during the first 6 seconds—when both cell-types were unilaterally in-phase—and z-scores relative to this early epoch were compared across the patient data and model for each motif-class and epoch. In each, the patient data and model were significantly correlated (*R, R*^2^ and raw *P*-values inset; maximal *P* after Benjamini-Hochberg-correction = 0.03). (**b**) The circular phase distances between the excitatory and inhibitory populations firing rates through time show a significant correlation between the model and the patient recording (*r* = 0.52; *P* < 0.001). (**c**) The mean firing rates for (***i***) the inhibitory and (***ii***) the excitatory populations, in response to adjusting the cross-synaptic weights. The inhibitory population firing rate is strongly affected by the strength of *w*_*IE*_, rapidly decreasing as the excitatory weighting increases. The excitatory population firing rate is mainly dependent on the strength of *w_IE_*. (**d**) Representative snapshots of various combinations of synaptic weights as labeled in **c**. Panel **d *iii*** is the combination of synaptic weights used in Fig. 7.

Cross correlation of the cell-type specific firing phases (i.e., the circular distance between excitatory and inhibitory phase angles through time, see Methods) showed significant similarity in the temporal evolution of excitatory and inhibitory firing patterns between the model and observed patient data (Fig. 8b; *r* = 0.52, *P* < 0.001), albeit with a small temporal shift of 0.11 s. In combination these show quantitative agreement between the model and patient data within individual motif-classes, across the full network, and with cell-type specific firing patterns.

We then sought to measure the inhibitory and excitatory network activities in response to adjusting the strength of the synaptic connections between these two networks (Fig. 8c, d). Four synaptic connection types exist in the model, excitatory onto excitatory and inhibitory cells (*w_EE_* and *w*_*IE*_ respectively), and inhibitory onto excitatory and inhibitory cells (*w_IE_* and *w_II_* respectively). Evaluating the mean firing rate in response to changes in the cross-synaptic weights revealed that strengthening *w*_*IE*_ results in a sharp decrease in inhibitory population firing rate in the 5 seconds preceding recruitment (Fig. 8c*i*). As expected, an increase in *w*_IE_ resulted in a reduction of excitatory activity (Fig. 8c*ii*). Similar out-of-phase inhibitory firing in the pre-recruitment period could be elicited across a range of cross-synaptic weightings, with subtle alterations to the onset speed and magnitude of inhibitory collapse (Fig. 8d). This analysis suggests this mechanism of delayed interneuron burst firing may commonly contribute independently to the inhibitory failure that permits seizure propagation.

## Discussion

Our findings highlight that spontaneous focal seizures in human neocortex show stereotyped inhibitory activity, with large increases in fast-spiking (FS) interneuronal firing early in the seizure. These increases, however, were delayed from the first ictal activity (“global” seizure onset), instead aligning to the moments immediately preceding the local tissue becoming “recruited” to the seizure-driving territory. This pre-recruitment activity involved a transition to tonic firing in the FS interneuron population, followed by a collapse in firing rate preceding the moment of maximal excitatory firing. Combining quantitative analysis of single neuronal firing patterns and computational modeling revealed repetitive, *transient* depolarization block as the likely culprit of this inhibitory firing reduction, and by extension, a likely mechanism of seizure propagation—especially into tissue that is not itself pathological.

This temporal relationship between cell-type specific firing and local recruitment becomes clearer when contextualized by the dual territory hypothesis of an ictal “core” of recruited tissue and a surrounding penumbra dominated by feedforward inhibition^22,26,27,62^. In this setting, ictal discharges emanate from a propagating wavefront through the cortex^45^, causing a band of increased inhibitory firing ahead of it, primarily (though not exclusively) via the fast-spiking, parvalbumin containing (PV+) interneurons^25,35^. This inhibitory firing must then collapse, as recently observed in an *in vivo* rodent model^63^, or become incapable of controlling runaway excitatory firing prior to that region being successfully recruited into the core, seizure-driving territory.

We showed previously that upon recruitment to this ictal core territory, which occurs on a cell-by-cell basis^28^ within cortical columns^22^, a neuron’s action potentials undergo waveform alterations due to the paroxysmal depolarizing shift, and as a result, the ictal wavefront can be defined by the local population’s action potential durations^28,44^. Analyzing the fast-spiking interneuronal firing rates as a function of this metric of local recruitment revealed that as the ictal wavefront approaches, the local inhibitory activity does indeed increase substantially, before subsiding (albeit still remaining above pre-ictal firing rates) as the tissue becomes recruited, in keeping with the prior temporal analyses (Fig. 3, Supplementary Movies 1 & 2).

The observed increase in inhibitory firing—immediately preceding ictal invasion—could be viewed as evidence that this interneuronal activity is in some way causative of the seizure^64^, as has been suggested for low-voltage fast activity onset seizures in human mesial structures^16^ and animal models^15,29^. The increases in this dataset of rhythmic onset neocortical seizures, however, appear instead to be attributable to the strong excitation derived from the nearby seizure^50^—the distribution of out-of-phase inhibitory firing across all 18 seizures from the 8 patients with rhythmic onset seizures makes it unlikely for the strong interneuron firing to be driving the seizure (Figs. 3 & 4). Moreover, both inhibitory and excitatory firing is at first heavily entrained to the rhythm after seizure onset, indicative that the shift to out-of-phase, followed by tonic firing, is driven from an unrecorded location elsewhere. These data are in keeping with recent *in vivo* work that suggested that, while depolarizing GABA may occur, it is rare outside of the experimental conditions used in *in vitro* work^65^.

Nevertheless, this does not exclude an interneuronal cause of any initial discharge that seeds the seizure itself, instead focusing on the method of ictal spread once the seizure is already underway (though the same mechanisms explored here would equally be viable to elucidate onset mechanisms, e.g. the “herald spike” at the start of low-voltage fast activity onset seizures eliciting a similar inhibitory response). The chloride-loading hypothesis, for example, is still feasible as a seizure initiation mechanism due to “static” chloride dysregulation in cells with decreased expression of the potassium-chloride co-transporter, KCC2^2,7,10,66,67^, even if it appears unlikely to be the primary cause of inhibitory failure as seizures spread through the neocortex, since this should result in peaks in excitatory firing concomitant with the inhibitory activity. Likewise, it is not improbable that once the seizure has spread to a region, i.e. after the reduction in inhibitory firing seen here, that chloride-loading in the pyramidal population occurs, further damaging the region’s inhibitory control. Optogenetic stimulation of PV+ interneurons in an *in vivo* model confirmed the plausibility of this scenario, showing a shift from anti- to pro-epileptic effects once the seizure was already underway, which could be suppressed via over-expression of KCC2^68^.

This focus here on ictal *propagation* mechanisms, rather than *onset* causes, might therefore seem relatively narrow in scope. However, many potential mechanisms for seizure onset have been suggested^12,69–71^ and it is likely that some, if not many, of these coexist. Acknowledging the constraints of seizure type and recording location, our data here suggest that propagation mechanisms, meanwhile, are likely more similar from patient to patient and seizure to seizure. Moreover, considering seizures have been hypothesized to originate from volumes smaller than 1 cubic mm^69,72^, it would appear doubtful that an electrode would often be sampling the true origin of the seizure, and so the majority of our mechanistic understanding *in vivo* is derived from propagated activity. Since the spread mechanism appears to be common across a variety of seizure onset patterns, targeting it in order to prevent the interneuronal out-of-phase firing and tonic transition may be broadly applicable.

During the propagation in these seizures, therefore, the question remains as to why the inhibition eventually gives way to allow the pathological activity to spread: what causes the firing rate reductions seen in the interneuronal population? One possibility is that these inhibitory cells are themselves being inhibited, for example by VIP+ cells^73^; a group that at present is not readily isolable from the population in extracellular recordings. Another plausible cause is a depletion of GABA vesicles in these highly active interneurons^74^.

An alternative explanation is that these interneurons are entering depolarization block, as has been suggested from *in vitro* models^24,32^, although studies have typically considered depolarization block as long-lasting (e.g. 5–40 seconds^32^), rather than repetitive, transient events, which would preclude their identification in these recordings since the cells overwhelmingly do not cease firing for prolonged durations. Continuously assessing action potential shapes, however, revealed amplitude fluctuations paired to the rhythmic, out-of-phase discharges in keeping with paroxysmal depolarizations followed by brief recovery (Fig. 6). Linear regression fits to these amplitudes through time, both within each burst and across consecutive bursts, revealed a stable decrease during the bursts at the population level, and in the majority of individual rhythmic onset seizures (15 out of 18; black dotted lines, Fig. 6c, d). Similarly, the inter-burst fits showed reductions in amplitude recovery between bursts at the population level, and in all but one rhythmic onset seizure (blue dashed lines, Fig. 6c, d), potentially indicating an increasing inability to recover between subsequent ictal discharges, leading to the eventual inhibitory failure and propagation of the seizure.

We hypothesized that this transient depolarization block was a result of the inhibitory interneurons’ smaller cell bodies^37^, and that their firing out-of-phase to the dominant ictal rhythm was a result of these blocks and recovery periods. To examine this, we used an intentionally uncomplicated Hodgkin-Huxley model^58^, to assess whether the firing properties are intrinsic to the fundamental properties of the two cell types. A simple arrangement of “resonators” for inhibitory cells and “integrators” for excitatory cells— based on previously established parameter sets^60^—was readily able to produce the firing patterns seen in the patient data, across a range of parameters (Figs. 7 & 8). Calculating the pseudo-extracellular trace for the modeled population by cell-type, and incorporating a threshold as used in spike sorting, revealed the transition to out-of-phase inhibitory activity was a result of saturation of the smaller cell-bodied interneurons. Without any intervention, this saturation was maximal during the peak of the dominant ictal rhythm, causing the action potentials to become subthreshold for “extracellular” detection, before recovering between bursts, but doing so less effectively as time progressed, mirroring accurately the human data observations (c.f. Fig. 6).

Taken together, these results suggest an intrinsic property of inhibitory interneurons gives rise to their eventual failure to restrain focal neocortical seizures. This mechanism does not require pathological connections between neurons, nor a weakening of their inhibitory efficacy downstream (though both may contribute also). Therefore, at its most basic level, neocortex is vulnerable to spreading, runaway excitation due to inhibitory interneurons’ predisposition to entering depolarization block transiently in the presence of repeated, intense excitation. As a result, therapies aimed—somewhat counterintuitively—at briefly hyperpolarizing interneurons, such as has recently been explored in an organotypic slice preparation^75^, could be a promising avenue for future interventions in neocortical focal epilepsy.

## Methods

### Human microelectrode recordings

Thirteen adult patients undergoing surgical evaluation for pharmacoresistant focal epilepsy across three clinical centers were implanted with “Utah”-style micro-electrode arrays (MEAs; Blackrock Microsystems, Salt Lake City, UT) simultaneous to standard clinical electrocorticography (ECoG). Informed consent was given by all participants prior to surgery and all procedures were approved by the respective Institutional Review Boards of Columbia University Medical Center, University of Utah and Massachusetts General Hospital/Brigham & Women’s Hospital. Clinical determination of the seizure onset zone (SOZ) and regions of spread were made by the treating physicians. MEAs were implanted into neocortical gyri based on presurgical estimation of the ictogenic region and consisted of 96 electrodes arranged in a 10 x 10 grid (with inactive corners) with an inter-electrode distance of 400 µm. Electrode lengths were either 1.0 mm (patients 1–10 & 13) or 1.5 mm (patients 11 & 12).

Neural data from the MEA were recorded at 30 kHz sample rate with a range of ± 8 mV with 16-bit precision, with a hardware filter between 0.3 Hz and 7.5 kHz. The reference was either subdural or epidural, chosen depending on recording quality. Simultaneous ECoG signals were recorded with sample rates of either 500 Hz or 2 kHz and a bandpass filter of 0.5 Hz to ¼ the sampling rate.

### Single-unit isolation and tracking

To account for waveshape alterations as a result of pathological activity and increases in background neural firing obscuring clusters, single-units were tracked through the seizure using convex hull-based template-matching, as previously described^28^.

Briefly, data from the peri-ictal period were bandpass filtered between 300 Hz and 5 kHz with a 1,024-order symmetric FIR filter to produce multi-unit activity (MUA) signals. The ictal period was blanked in order to perform initial spike sorting only on stable, baseline data,^44^ then spikes were detected with a threshold of 4.5 times the standard deviation as estimated from the median absolute deviation^76^. Spikes were clustered on a channel-by-channel basis using a modified version of the “UltraMegaSort2000” MATLAB toolbox^77–79^. Clusters were deemed single units if they satisfied the following criteria: (a) clear separation from other clusters on Fisher’s linear discriminant in principal component space; (b) <1% spikes within the 2 ms absolute refractory period; (c) absence of outliers based on the expected ξ^2^ distribution of Mahalanobis distances; and (d) <1% missing spikes below threshold for detection as estimated by a Gaussian distribution fit to spike voltages^78^.

Waveforms were then detected in the full peri-ictal period, including the seizure, in a similar manner though without clustering. Principal component scores were calculated for these spikes based on the principal component space defined during the original spike sorting. Waveforms that fell within the convex hull of a previously defined single unit were selected as putative action potentials from the same neuron. The confidence that each spike arose from its assigned neuron was then calculated based on Gaussian fits for the voltage at each data point in the original single-unit. This method allows for more accurate tracking of neuronal firing despite action potential waveform alterations as a result of ion concentration changes or paroxysmal depolarization shifts, or due to an increase in “noise” levels obscuring previously isolated clusters^28^.

### Cell-type subclassification

The original single-units—prior to template matching for tracking through the seizure—were subclassified into putative cell-types based on mean extracellular waveforms and cell-intrinsic firing pattern autocorrelation as previously described^42^. Mean waveforms were calculated as the action potential-triggered average from the original, unfiltered signal and then z-scored. The unit’s firing rate autocorrelation over ± 50 ms was calculated in 0.5 ms bins.

Putative fast-spiking (FS) interneurons were isolated probabilistically from the regular-spiking population using a 2-component Gaussian mixture model on each unit’s mean waveforms in principal component space (Fig. 1a), a method which has been shown to separate inhibitory FS cells from the regular spiking population^80^.

To isolate putative inhibitory interneurons from the remaining non-FS population, a set of exponential equations was fitted to the autocorrelation for each unit not already classified as an FS interneuron, using the “fit_ACG” function from the “CellExplorer” MATLAB toolbox^81^. To calculate the probability of a unit being inhibitory or excitatory we fitted a 2-component Gaussian mixture model to the *τ*_*rise*_ exponential, which captures the cell-intrinsic propensity to fire bursts of action potentials—a feature that has been shown to separate putative excitatory and inhibitory cells^81–83^ (Fig. 1b).

### Ictal recruitment and probabilistic firing rates

A region defined by intense synaptic activity—due to the strong excitatory drive from the seizure—but without entrained, aberrant neuronal firing due to feedforward inhibition exists within tissue prior to its involvement in driving a seizure^22,23,26,62^. The moment of this transition (“ictal recruitment”) was defined at each microelectrode as previously described^28,45^. Briefly, the hallmark of ictal recruitment is transient, tonic neuronal firing, which subsequently transitions to “burst” firing as the wavefront of recruitment propagates away from the region, seeding rhythmic discharges back into the area^45,56^. To quantitatively define this moment of recruitment, a Gaussian kernel with 200 ms SD was convolved with the spike times detected in the MUA. A sustained, significant increase in this signal followed by a transition to burst firing was determined as the moment of local recruitment^45^. Ictal recordings without this signature activity were classified as unrecruited. Analyses of the “pre-recruitment” period for the whole MEA were calculated on an epoch between “global” seizure onset and the mean + SD of the wavefront time calculated across all channels on the MEA. Note that “pre-recruitment” refers to the time prior to local recruitment but after global seizure onset, rather than to a specific state of being “pre-recruited”.

Template-matching with convex hulls is intentionally permissive, to avoid undercounting activity from neurons whose action potential shapes have been altered or dropped below the noise threshold^28^. To account for this, instantaneous firing rates were calculated by convolving each unit’s spike times with a Gaussian kernel that was scaled to the confidence that that spike had arisen from its assigned unit, as described above and in more detail previously^28^. All subsequent analyses of firing times and patterns were weighted by the confidence for each action potential of interest. Z-scored firing rate changes within neurons (Fig. 3b) were calculated via the standard deviation of a Poisson distribution with the observed firing rates^42,84^.

### Phase analyses in rhythmic onset seizures

Further analysis was performed on seizures with rhythmic onsets: to account for fluctuations in discharge timings, analyses of firing patterns were performed with respect to the instantaneous phase of the dominant ictal rhythm. Entrainment was calculated using the Rayleigh *z*-test, and unless specified otherwise, statistical differences in phase angles were calculated with the circular Kuiper test^85^.

Spiral time-phase plots were calculated by organizing the firing times of every neuron by their phase angle with respect to the dominant ictal rhythm through time, starting with global seizure onset at the center, and ending with local recruitment at the outer edge of the spiral (Fig. 4c*i*). As a result, ictal time— with respect to the instantaneous phase angle of the dominant ictal rhythm—is shown progressively expanding counter-clockwise from the center of the spiral until ictal recruitment at the outer edge. The gaussian-convolved (SD = 20 ms) firing rates could then be calculated as described above for each cell-type population, and smoothed on the resultant spiral (angular smoothing of 30°, radial smoothing of 0.1) to give an instantaneous estimate of phase-specific excitatory versus inhibitory firing evolving through time (Fig. 4c*ii*, Fig. 5c).

Spike amplitude trajectories through time (Fig. 6) were calculated as linear regressions both within each burst, and as an overall fit through the pre-recruitment epoch, thus resulting in a two-dimensional plane for each seizure. Fits during the intra-burst time was limited to the lesser of 500 ms or ¾ of the duration until the next discharge’s peak, to avoid capturing the onset of the following discharge or analyzing firing unrelated to the current discharge.

### Hodgkin–Huxley model

We simulated network activity of cell-type interactions during the transition from seizure onset to recruitment via a 10-neuron cortical model characterized by Hodgkin-Huxley type dynamics, in which the membrane potential, *V*, is governed by the following equation^58^:

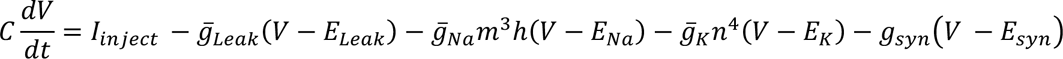

where *I* is current, *C* is the membrane capacitance, 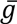 is the maximal conductance, and *m*, *n*, and *h* are dimensionless variables associated with activation for sodium and potassium channels and inactivation for sodium channels respectively. Synaptic conductance, *g_syn_*, is modeled by the alpha function: ∝ *βte^−∝βt^* for *t* ≥ 0, where ∝ and *β* are constants specific to cell-type synapses^37,86,87^. As per Tryba et al.^37^, for an excitatory synapse, ∝ = 25 and β = 0.3 ms^−1^; for an inhibitory synapse, ∝ = 3 and β = 0.1 ms^−1^.

Using previously established parameter sets^60^, we implemented fast-spiking/resonator (“Type II”^88,89^) behavior for the inhibitory cell type and regular-spiking/integrator (“Type I”^88,89^) activity for the excitatory cell type (Fig. 7a, Tables S2 & S3). These cell-types were configured in a network with 4:1 ratio for excitatory and inhibitory neurons respectively (Fig. 7b). The cortical model was placed in a passing ictal wave and rhythm (*I_inject_*), modeled as an exponential growth function and sinusoidal waveform respectively, and the resulting activity was determined (Fig. 7c). Assuming an extracellular electrode equidistant from the model neurons, its signal was determined as the signal proportional to the sum of the second derivatives of all membrane potentials, representing the transmembrane currents^90^.

To quantify similarities between the model outputs and the observed patient data, we computed triple correlation^61^ for the resultant spike raster both for the model and for a representative patient recording (Patient 3, seizure 1; Fig. 4a; each sampled at 500 Hz). Triple correlation is a method that relates three nodes: one reference node and up to two other nodes separated by up to two lags in both space (*n*_1_ & *n*_2_) and time (*t*_1_ & *t*_2_)^61^. These three-node configurations can be collapsed into fourteen qualitatively distinct motif classes. Furthermore, these third-order motif configurations are sufficient for a complete and unique characterization of the network activity^61^. Triple correlation was calculated in 1 second bins, with a spatial window that covered the entirety of the dataset and a temporal window of ±50 ms. To compare firing phases between the model and patient data, cell-type specific population Gaussian-convolved instantaneous firing rates (see above) in each were de-meaned and narrow-band filtered (3– 5.5 Hz; 500^th^ order symmetric FIR), and their instantaneous phase angles calculated using the Hilbert transform. Cross correlation was then calculated on the circular distance between the two cell-types’ phase angles was then calculated through time both for the model and the patient data.

## Data availability

The raw data that support the findings of this study are available upon reasonable request from the corresponding author. To protect the privacy of research participants, they are not publicly accessible.

## Code availability

Accompanying code is available at https://github.com/edmerix/

## Author contributions

E.M.M., E.H.S., E.D.S., G.M.M., R.R.G., S.A.S., B.G., P.A.H., E.N.E., J.R.M., S.S.C. and C.A.S. were involved in collecting the data. E.M.M., S.S.D., E.H.S., A.J.T., W.v.D and C.A.S. conceived the study. E.M.M., A.H.A-M. and E.H.S. analyzed the data. S.S.D. and W.v.D. created the computational model. E.M.M., S.S.D., B.G., A.J.T., W.v.D. and C.A.S. drafted and edited the manuscript. All authors approved the final version.

## Funding

R01 NS084142 and R01 NS110669 (C.A.S.), University of Chicago MSTP Training Grant T32GM007281 and F31NS127493 (S.S.D.).

## Competing interests

S.A.S. is a consultant for Boston Scientific, Zimmer Biomet, Neuropace, Koh Young, Sensoria Therapeutics and Varian Medical, and is co-founder of Motif Neurotech. The other authors report no competing interests.

## Notes

### Funding Statement

The authors acknowledge funding from R01 NS084142 and R01 NS110669 (C.A.S.), and University of Chicago MSTP Training Grant T32GM007281 and F31NS127493 (S.S.D.)

### Author Declarations

All procedures were approved by the respective Institutional Review Boards of Columbia University Medical Center, University of Utah and Massachusetts General Hospital/Brigham & Women’s Hospital.

## References

1. Moshé, S. L., Perucca, E., Ryvlin, P. & Tomson, T. Epilepsy: new advances. Lancet Lond. Engl. 385, 884–898 (2015).

2. Weiss, S. A. Chloride ion dysregulation in epileptogenic neuronal networks. Neurobiol. Dis. 177, 106000 (2023).

3. Wenzel, M., Huberfeld, G., Grayden, D. B., de Curtis, M. & Trevelyan, A. J. A debate on the neuronal origin of focal seizures. Epilepsia (2023) doi:10.1111/epi.17650.

4. Graham, R. T. et al. Optogenetic stimulation reveals a latent tipping point in cortical networks during ictogenesis. Brain J. Neurol. 146, 2814–2827 (2023).

5. Trevelyan, A. J., Graham, R. T., Parrish, R. R. & Codadu, N. K. Synergistic Positive Feedback Mechanisms Underlying Seizure Initiation. Epilepsy Curr. 23, 38–43 (2023).

6. Dzhala, V. I. et al. Progressive NKCC1-dependent neuronal chloride accumulation during neonatal seizures. J. Neurosci. 30, 11745–11761 (2010).

7. Huberfeld, G. et al. Perturbed chloride homeostasis and GABAergic signaling in human temporal lobe epilepsy. J. Neurosci. 27, 9866–9873 (2007).

8. Lillis, K. P., Kramer, M. A., Mertz, J., Staley, K. J. & White, J. A. Pyramidal cells accumulate chloride at seizure onset. Neurobiol. Dis. 47, 358–366 (2012).

9. Alfonsa, H. et al. The Contribution of Raised Intraneuronal Chloride to Epileptic Network Activity. J. Neurosci. 35, 7715–7726 (2015).

10. Pallud, J. et al. Cortical GABAergic excitation contributes to epileptic activities around human glioma. Sci. Transl. Med. 6, 244ra89 (2014).

11. Ellender, T. J., Raimondo, J. V., Irkle, A., Lamsa, K. P. & Akerman, C. J. Excitatory Effects of Parvalbumin-Expressing Interneurons Maintain Hippocampal Epileptiform Activity via Synchronous Afterdischarges. J. Neurosci. 34, 15208–15222 (2014).

12. Bragin, A., Wilson, C. L. & Engel, J. Chronic Epileptogenesis Requires Development of a Network of Pathologically Interconnected Neuron Clusters: A Hypothesis. Epilepsia 41, S144–S152 (2000).

13. Yang, J. C. et al. Microscale dynamics of electrophysiological markers of epilepsy. Clin. Neurophysiol. S1388245721006702 (2021) doi:10.1016/j.clinph.2021.06.024.

14. Chang, M. et al. Brief activation of GABAergic interneurons initiates the transition to ictal events through post-inhibitory rebound excitation. Neurobiol. Dis. 109, 102–116 (2018).

15. de Curtis, M. & Avoli, M. GABAergic networks jump-start focal seizures. Epilepsia 57, 679–687 (2016).

16. Elahian, B. et al. Low-voltage fast seizures in humans begin with increased interneuron firing. Ann. Neurol. 84, 588–600 (2018).

17. Prince, D. A. & Wilder, B. J. Control mechanisms in cortical epileptogenic foci. ‘Surround’ inhibition. Arch Neurol 16, 194–202 (1967).

18. Dichter, M. & Spencer, W. A. Penicillin-induced interictal discharges from the cat hippocampus. II. Mechanisms underlying origin and restriction. J Neurophysiol 32, 663–687 (1969).

19. Wong, B. Y. & Prince, D. A. The lateral spread of ictal discharges in neocortical brain slices. Epilepsy Res 7, 29–39 (1990).

20. Schwartz, T. H. & Bonhoeffer, T. In vivo optical mapping of epileptic foci and surround inhibition in ferret cerebral cortex. Nat Med 7, 1063–1067 (2001).

21. Timofeev, I., Grenier, F. & Steriade, M. The role of chloride-dependent inhibition and the activity of fast-spiking neurons during cortical spike-wave electrographic seizures. Neuroscience 114, 1115–1132 (2002).

22. Trevelyan, A. J., Sussillo, D., Watson, B. O. & Yuste, R. Modular propagation of epileptiform activity: evidence for an inhibitory veto in neocortex. J. Neurosci. 26, 12447–12455 (2006).

23. Trevelyan, A. J., Sussillo, D. & Yuste, R. Feedforward inhibition contributes to the control of epileptiform propagation speed. J. Neurosci. 27, 3383–3387 (2007).

24. Cammarota, M., Losi, G., Chiavegato, A., Zonta, M. & Carmignoto, G. Fast spiking interneuron control of seizure propagation in a cortical slice model of focal epilepsy. J. Physiol. 591, 807–822 (2013).

25. Parrish, R. R., Codadu, N. K., Mackenzie Gray Scott, C. & Trevelyan, A. J. Feedforward inhibition ahead of ictal wavefronts is provided by both parvalbumin- and somatostatin-expressing interneurons. J. Physiol. 597, 2297–2314 (2019).

26. Schevon, C. A. et al. Evidence of an inhibitory restraint of seizure activity in humans. Nat. Commun. 3, 1060–11 (2012).

27. Weiss, S. A. et al. Ictal high frequency oscillations distinguish two types of seizure territories in humans. Brain 136, 3796–3808 (2013).

28. Merricks, E. M. et al. Neuronal Firing and Waveform Alterations through Ictal Recruitment in Humans. J. Neurosci. 41, 766–779 (2021).

29. Gnatkovsky, V., Librizzi, L., Trombin, F. & de Curtis, M. Fast activity at seizure onset is mediated by inhibitory circuits in the entorhinal cortex in vitro. Ann. Neurol. 64, 674–686 (2008).

30. Shiri, Z., Manseau, F., Lévesque, M., Williams, S. & Avoli, M. Interneuron activity leads to initiation of low-voltage fast-onset seizures: Epileptiform Synchronization. Ann. Neurol. 77, 541–546 (2015).

31. Lévesque, M., Herrington, R., Hamidi, S. & Avoli, M. Interneurons spark seizure-like activity in the entorhinal cortex. Neurobiol. Dis. 87, 91–101 (2016).

32. Ziburkus, J., Cressman, J. R., Barreto, E. & Schiff, S. J. Interneuron and pyramidal cell interplay during in vitro seizure-like events. J. Neurophysiol. 95, 3948–3954 (2006).

33. Misra, A., Long, X., Sperling, M. R., Sharan, A. D. & Moxon, K. A. Increased neuronal synchrony prepares mesial temporal networks for seizures of neocortical origin. Epilepsia 3, 219–14 (2018).

34. Trevelyan, A. J. & Schevon, C. A. How inhibition influences seizure propagation. Neuropharmacology 69, 45–54 (2013).

35. Sessolo, M. et al. Parvalbumin-Positive Inhibitory Interneurons Oppose Propagation But Favor Generation of Focal Epileptiform Activity. J. Neurosci. 35, 9544–9557 (2015).

36. Bikson, M., Hahn, P. J., Fox, J. E. & Jefferys, J. G. R. Depolarization block of neurons during maintenance of electrographic seizures. J. Neurophysiol. 90, 2402–2408 (2003).

37. Tryba, A. K. et al. Role of paroxysmal depolarization in focal seizure activity. J. Neurophysiol. 122, 1861–1873 (2019).

38. Huberfeld, G. et al. Glutamatergic pre-ictal discharges emerge at the transition to seizure in human epilepsy. Nat. Neurosci. 14, 627–634 (2011).

39. Viitanen, T., Ruusuvuori, E., Kaila, K. & Voipio, J. The K ^+^ -Cl ^−^ cotransporter KCC2 promotes GABAergic excitation in the mature rat hippocampus: GABA excitation and KCC2. J. Physiol. 588, 1527–1540 (2010).

40. Köhling, R., D’Antuono, M., Benini, R., de Guzman, P. & Avoli, M. Hypersynchronous ictal onset in the perirhinal cortex results from dynamic weakening in inhibition. Neurobiol. Dis. 87, 1–10 (2016).

41. Miri, M. L., Vinck, M., Pant, R. & Cardin, J. A. Altered hippocampal interneuron activity precedes ictal onset. eLife 7, 1277 (2018).

42. Agopyan-Miu, A. H. et al. Cell-type specific and multiscale dynamics of human focal seizures in limbic structures. Brain 146, 5209–5223 (2023).

43. Buzsàki, G., Anastassiou, C. A. & Koch, C. The origin of extracellular fields and currents — EEG, ECoG, LFP and spikes. Nat. Rev. Neurosci. 13, 407–420 (2012).

44. Merricks, E. M. et al. Single unit action potentials in humans and the effect of seizure activity. Brain J. Neurol. 138, 2891–2906 (2015).

45. Smith, E. H. et al. The ictal wavefront is the spatiotemporal source of discharges during spontaneous human seizures. Nat Commun 7, 11098 (2016).

46. Diamond, J. M. et al. Travelling waves reveal a dynamic seizure source in human focal epilepsy. Brain 144, 1751–1763 (2021).

47. Schlafly, E. D. et al. Multiple Sources of Fast Traveling Waves during Human Seizures: Resolving a Controversy. J. Neurosci. 42, 6966–6982 (2022).

48. McCormick, D. A., Connors, B. W., Lighthall, J. W. & Prince, D. A. Comparative electrophysiology of pyramidal and sparsely spiny stellate neurons of the neocortex. J. Neurophysiol. 54, 782–806 (1985).

49. Barthó, P. et al. Characterization of neocortical principal cells and interneurons by network interactions and extracellular features. J. Neurophysiol. 92, 600–608 (2004).

50. Smith, E. H. et al. Dual mechanisms of ictal high frequency oscillations in human rhythmic onset seizures. Sci. Rep. 10, 19166 (2020).

51. Liou, J. et al. Role of inhibitory control in modulating focal seizure spread. Brain J. Neurol. 141, 2083–2097 (2018).

52. Liou, J. et al. A model for focal seizure onset, propagation, evolution, and progression. eLife 9, e50927 (2020).

53. Tremblay, R., Lee, S. & Rudy, B. GABAergic Interneurons in the Neocortex: From Cellular Properties to Circuits. Neuron 91, 260–292 (2016).

54. Kandel, E. R. & Spencer, W. A. The pyramidal cell during hippocampal seizure. Epilepsia 2, 63–69 (1961).

55. Kandel, E. R. & Spencer, W. A. Electrophysiology of hippocampal neurons. II. After-potentials and repetitive firing. J Neurophysiol 24, 243–259 (1961).

56. Matsumoto, H. & Ajmone Marsan, C. Cortical cellular phenomena in experimental epilepsy: Ictal manifestations. Exp Neurol 9, 305–326 (1964).

57. Traub, R. D. & Wong, R. K. Cellular mechanism of neuronal synchronization in epilepsy. Science 216, 745–747 (1982).

58. Hodgkin, A. L. & Huxley, A. F. A quantitative description of membrane current and its application to conduction and excitation in nerve. J. Physiol. 117, 500–544 (1952).

59. Paré, D., deCurtis, M. & Llinás, R. Role of the hippocampal-entorhinal loop in temporal lobe epilepsy: extra- and intracellular study in the isolated guinea pig brain in vitro. J. Neurosci. Off. J. Soc. Neurosci. 12, 1867–1881 (1992).

60. Bukoski, A., Steyn-Ross, D. A. & Steyn-Ross, M. L. Channel-noise-induced critical slowing in the subthreshold Hodgkin-Huxley neuron. Phys. Rev. E Stat. Nonlin. Soft Matter Phys. 91, 032708 (2015).

61. Deshpande, S. S., Smith, G. A. & Van Drongelen, W. Third-order motifs are sufficient to fully and uniquely characterize spatiotemporal neural network activity. Sci. Rep. 13, 238 (2023).

62. Schevon, C. A. et al. Multiscale recordings reveal the dynamic spatial structure of human seizures. Neurobiol. Dis. 127, 303–311 (2019).

63. Shimoda, Y. et al. Extracellular glutamate and GABA transients at the transition from interictal spiking to seizures. Brain awad336 (2023) doi:10.1093/brain/awad336.

64. Dossi, E. & Huberfeld, G. GABAergic circuits drive focal seizures. Neurobiol. Dis. 180, 106102 (2023).

65. Dubanet, O. et al. Probing the polarity of spontaneous perisomatic GABAergic synaptic transmission in the mouse CA3 circuit in vivo. Cell Rep. 36, 109381 (2021).

66. Chen, L. et al. KCC2 downregulation facilitates epileptic seizures. Sci. Rep. 7, 156 (2017).

67. Di Cristo, G., Awad, P. N., Hamidi, S. & Avoli, M. KCC2, epileptiform synchronization, and epileptic disorders. Prog. Neurobiol. 162, 1–16 (2018).

68. Magloire, V., Cornford, J., Lieb, A., Kullmann, D. M. & Pavlov, I. KCC2 overexpression prevents the paradoxical seizure-promoting action of somatic inhibition. Nat. Commun. 10, 1225 (2019).

69. Stead, M. et al. Microseizures and the spatiotemporal scales of human partial epilepsy. Brain 133, 2789–2797 (2010).

70. Wagner, F. B. et al. Microscale spatiotemporal dynamics during neocortical propagation of human focal seizures. NeuroImage 122, 114–130 (2015).

71. Wang, Y. et al. Mechanisms underlying different onset patterns of focal seizures. PLOS Comput. Biol. 13, e1005475 (2017).

72. Wenzel, M., Hamm, J. P., Peterka, D. S. & Yuste, R. Acute Focal Seizures Start As Local Synchronizations of Neuronal Ensembles. J. Neurosci. 39, 8562–8575 (2019).

73. Karnani, M. M., Agetsuma, M. & Yuste, R. A blanket of inhibition: functional inferences from dense inhibitory connectivity. Curr. Opin. Neurobiol. 26, 96–102 (2014).

74. Zhang, Z. J. et al. Transition to seizure: ictal discharge is preceded by exhausted presynaptic GABA release in the hippocampal CA3 region. J. Neurosci. 32, 2499–2512 (2012).

75. Călin, A., Ilie, A. S. & Akerman, C. J. Disrupting Epileptiform Activity by Preventing Parvalbumin Interneuron Depolarization Block. J. Neurosci. 41, 9452–9465 (2021).

76. Quian Quiroga, R., Nadasdy, Z. & Ben-Shaul, Y. Unsupervised Spike Detection and Sorting with Wavelets and Superparamagnetic Clustering. Neural Comput. 16, 1661–1687 (2004).

77. Fee, M. S., Mitra, P. P. & Kleinfeld, D. Automatic sorting of multiple unit neuronal signals in the presence of anisotropic and non-Gaussian variability. J. Neurosci. Methods 69, 175–188 (1996).

78. Hill, D. N., Mehta, S. B. & Kleinfeld, D. Quality metrics to accompany spike sorting of extracellular signals. J. Neurosci. 31, 8699–8705 (2011).

79. Merricks, E. edmerix/SplitMerge: v1.0. Zenodo 10.5281/zenodo.3951171 (2020).

80. Ardid, S. et al. Mapping of Functionally Characterized Cell Classes onto Canonical Circuit Operations in Primate Prefrontal Cortex. J. Neurosci. 35, 2975–2991 (2015).

81. Petersen, P. C., Siegle, J. H., Steinmetz, N. A., Mahallati, S. & Buzsáki, G. CellExplorer: A framework for visualizing and characterizing single neurons. Neuron 109, 3594–3608.e2 (2021).

82. Petersen, P. C. & Buzsáki, G. Cooling of Medial Septum Reveals Theta Phase Lag Coordination of Hippocampal Cell Assemblies. Neuron 107, 731–744.e3 (2020).

83. Senzai, Y. & Buzsáki, G. Physiological Properties and Behavioral Correlates of Hippocampal Granule Cells and Mossy Cells. Neuron 93, 691–704.e5 (2017).

84. Vajda, I. et al. Low-frequency stimulation induces stable transitions in stereotypical activity in cortical networks. Biophys. J. 94, 5028–5039 (2008).

85. Berens, P. CircStat: a MATLAB toolbox for circular statistics. J Stat Softw (2009).

86. Lopes Da Silva, F. H., Vos, J. E., Mooibroek, J. & Van Rotterdam, A. Relative contributions of intracortical and thalamo-cortical processes in the generation of alpha rhythms, revealed by partial coherence analysis. Electroencephalogr. Clin. Neurophysiol. 50, 449–456 (1980).

87. van Drongelen, W. Signal Processing for Neuroscientists. (Academic press, 2018).

88. Hodgkin, A. L. The local electric changes associated with repetitive action in a non-medullated axon. J. Physiol. 107, 165–181 (1948).

89. Izhikevich, E. M. Which model to use for cortical spiking neurons? IEEE Trans. Neural Netw. 15, 1063–1070 (2004).

90. Clark, J. & Plonsey, R. The extracellular potential field of the single active nerve fiber in a volume conductor. Biophys. J. 8, 842–864 (1968).

