## Supplementary Fig. 1 for "Aberrant fast spiking interneuronal activity precedes seizure transitions in humans"


**Contents**


- Supplementary Figure 1
- Table S1
- Table S2
- Table S3
- Supplementary Movie 1 legend
- Supplementary Movie 2 legend

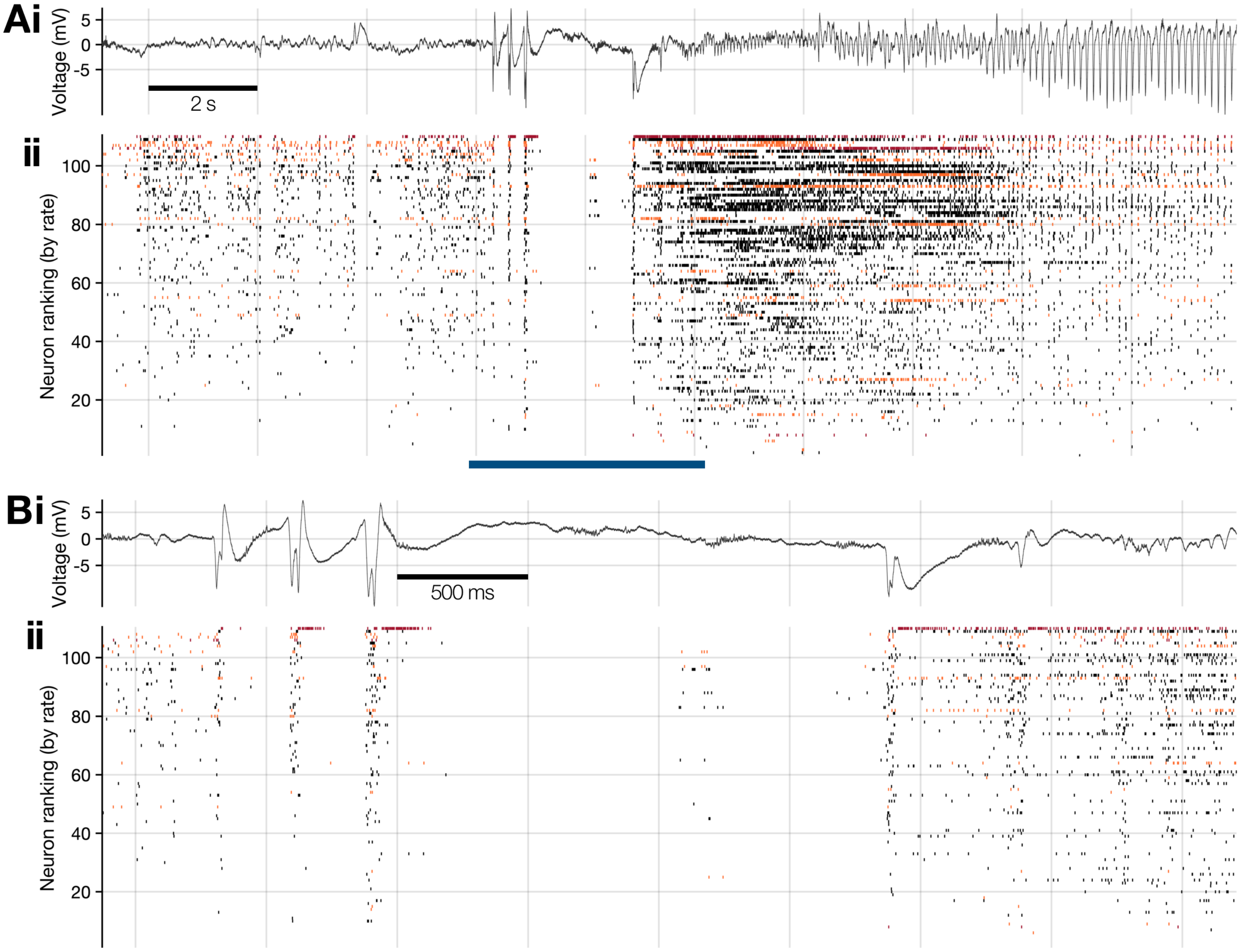


**Supplementary Figure 1. Neuronal firing underlying a spontaneous, human focal seizure. (a)(*i*)** Mean LFP calculated across the Utah array during patient 4's seizure, bandpass filtered between 2–50 Hz. **(*ii*)** Underlying, time-locked neuronal firing across the full Utah array, tracked with convex hulls[28], ordered by each neuron's firing rate during the epoch, with putative pyramidal cells in black, "wide" interneurons in orange, and FS interneurons in red. **(b)** Enlargement of the epoch marked with a blue bar in (a), using the same layout.

| Patient | Age range | Sex | MEA length (mm) | Implant location | Seizure onset | Pathology | Seizure # | Seizure type | Rhythmic onset | Evidence of recruitment | Total neurons | Total FS interneurons |
|---|---|---|---|---|---|---|---|---|---|---|---|---|
| 1 | 26–30 | M | 1.0 | Left frontal convexity | Left supplementary motor area | Nonspecific | 1 | FIA | - | N | 75 | 0 |
| | | | | | | | 2 | FIA | - | N | 75 | 0 |
| | | | | | | | 3 | FIA | - | N | 50 (-4) | 1 |
| 2 | 36–40 | M | 1.0 | Left dorsolateral frontal lobe | Left frontal operculum | Nonspecific | 1 | FIA | - | N | 61 | 3 |
| | | | | | | | 2 | FIA | - | N | 65 | 2 |
| | | | | | | | 3 | FIA | - | N | 59 (-2) | 0 |
| 3 | 31–35 | F | 1.0 | Left inferior temporal gyrus | Left basal/anterior temporal | Mild CA1 neuronal loss; lateral temporal nonspecific | 1 | FIA | Y | Y | 111 | 17 |
| | | | | | | | 2 | FIA | Y | Y | 101 | 3 |
| | | | | | | | 3 | FIA | Y | Y | 91 | 6 |
| 4 | 18–25 | F | 1.0 | Right posterior temporal gyrus | Right posterior lateral temporal | Nonspecific | 1 | FTBTC | - | Y | 110 | 1 |
| 5 | 26–30 | M | 1.0 | Left premotor | Left prefrontal | Mild reactive astrogliosis; patchy microgliosis; Chaslin's marginal sclerosis | 1 | FTBTC | Y | Y | 120 | 9 |
| | | | | | | | 2 | FTBTC | - | Y | 309 | 15 |
| 6 | 26–30 | M | 1.0 | Left posterior inferior temporal gyrus | Left subtemporal/lateral temporal | Diffusely infiltrating low grade glioma, IDH-1 negative | 1 | FIA | Y | Y | 165 | 9 |
| | | | | | | | 2 | FIA | Y | Y | 104 | 10 |
| | | | | | | | 3 | FIA | - | Y | 102 | 11 |
| 7 | 26–30 | M | 1.0 | Right mesial temporal gyrus | Right subtemporal | Mild astrocytosis | 1 | FA | - | Y (partial) | 240 | 19 |
| | | | | | | | 2 | FA | - | Y | 252 (-16) | 13 (-2) |
| | | | | | | | 3 | FA | - | Y | 241 | 14 |
| 8 | 26–30 | M | 1.0 | Left anterior/inferior temporal | Left frontal polar/orbitofrontal | Focal cortical dysplasia type 2a; Reactive astrogliosis | 1 | FIA | - | Y | 77 | 7 |
| | | | | | | | 2 | FTBTC | - | Y | 78 | 11 |
| | | | | | | | 3 | FTBTC | - | Y | 77 | 1 |
| 9 | 18–25 | M | 1.0 | Left inferior temporal gyrus | Left mesial temporal with spread to lateral temporal | Mesial temporal sclerosis; nonspecific | 1 | FIA | Y | Y | 119 (-1) | 9 |
| | | | | | | | 2 | FIA | Y | Y | 133 | 14 |
| 10 | 31–35 | M | 1.0 | Right inferior frontal gyrus | Right mesial temporal | Nonspecific | 1 | FTBTC | Y | Y | 86 | 4 |
| 11 | 41–45 | M | 1.5 | Right middle temporal gyrus | Right temporal | Mesial temporal sclerosis | 1 | FTBTC | Y | Y | 109 | 10 |
| | | | | | | | 2 | FTBTC | Y | Y | 83 | 5 |
| | | | | | | | 3 | FTBTC | Y | Y | 137 | 7 |
| 12 | 31–35 | M | 1.5 | Left middle temporal gyrus | Mesial temporal | Mesial temporal sclerosis | 1 | FIA | Y | Y | 234 | 11 |
| | | | | | | | 2 | FIA | Y | Y | 189 | 10 |
| | | | | | | | 3 | FIA | Y | Y | 230 | 14 |
| 13 | 18–25 | M | 1.0 | Left superior temporal gyrus | Left temporal | Mesial temporal sclerosis | 1 | FIA | Y | Y | 121 | 10 |
| | | | | | | | 2 | FIA | Y | Y | 91 | 5 |
| | | | | | | | 3 | FIA | Y | Y | 104 | 5 |

**Table S1. Patient demographics.** Numbers in parentheses represent total neurons that were not trackable during the seizure via convex hull template matching[28]. FTBTC = focal to bilateral tonic clonic; FIA = focal with impaired awareness; FA = focal aware.

**Cellular parameters**

| | Resonator | Integrator | Units |
|---|---|---|---|
| **Capacitance** | 1 | 3 | $\mu F/cm^2$ |
| $\mathbf{E_{Na}}$ | 55 | 50 | mV |
| $\mathbf{E_K}$ | -72 | -95 | mV |
| $\mathbf{E_L}$ | -49.4 | -63.7 | mV |
| $\mathbf{g_{Na}}$ | 120 | 25 | $mS/cm^2$ |
| $\mathbf{g_K}$ | 36 | 5 | $mS/cm^2$ |
| $\mathbf{g_L}$ | 0.3 | 0.1 | $mS/cm^2$ |

**Table S2. Cellular parameters used in Hodgkin-Huxley model.** Values reproduced/scaled from Bukoski *et al.*[60]

| Symbol | Resonator | Integrator |
|---|---|---|
| $\alpha_n(V)$ | $0.01(V + 50)/\{1 - \exp[-(V + 50)/10]\}$ | $-0.032(V + 50)/\{\exp[-(V + 50)/5] - 1\}$ |
| $\alpha_m(V)$ | $0.1(V + 35)/\{1 - \exp[-(V + 35)/10]\}$ | $-0.0053(V + 52)/\{\exp[-(V + 52)/4] - 1\}$ |
| $\alpha_h(V)$ | $0.07 \exp[-(V + 60)/20]$ | $0.128 \exp[-(V + 48)/18]$ |
| $\beta_n(V)$ | $0.125 \exp[-(V + 60)/80]$ | $0.084 \exp[-(V + 55)/40]$ |
| $\beta_m(V)$ | $4 \exp[-(V + 60)/18]$ | $0.28(V + 25)/\{\exp[(V + 25)/5] - 1\}$ |
| $\beta_h(V)$ | $1/\{\exp[-(V + 30)/10] + 1\}$ | $4/\{\exp[-(V + 25)/5] + 1\}$ |

**Table S3. Gating equations for resonator and integrator classes.** Reproduced/scaled from Bukoski *et al.*[60]

### Supplementary Movie Legends:

**Supplementary Movie 1. Fast-spiking interneuron activity during ictal propagation, with successful inhibitory restraint.** *Above*: Mean LFP from the Utah array during patient 7's first seizure, with "global" onset and current time marked (blue triangle and red line respectively). *Below*: A heatmap showing the 2-dimensional linear regression fit to all excitatory cells' spike full-width at half-maximum (FWHM; z-axis and heatmap color) with respect to their location on the Utah array (x- and y-axes), at each moment in time (5 s window, advancing in 100 ms steps). Each red dot embedded in the plane represents one fast-spiking (FS) interneuron, with the diameter showing that cell's instantaneous firing rate. Note the increase in inhibitory activity across the array shortly after global seizure onset (~15 seconds through movie), the approaching ictal wavefront at the nearest corner (x = 3.6 mm, y = 0.0 mm; ~27 seconds; represented by an increase in the excitatory population FWHM), followed by sudden reductions in nearby FS interneuron firing rates ahead of the ictal wavefront propagating into that region (~30 seconds till end). Note only half the array was successfully recruited to the seizure before termination, with maintained FS inhibitory firing outside the recruited region (c.f. Fig. 2 in main text).

**Supplementary Movie 2. Fast-spiking interneuron activity during ictal propagation across whole array.** Same design and layout as Supplementary Movie 1, using patient 7's second seizure, where the seizure managed to spread across the full Utah array prior to ictal termination. Note the increasing FS interneuron firing rates just prior to the excitatory population's FWHM increase, and their collapse as the local excitatory population FWHM reaches its peak at each location, with a temporal progression from right to left.
